# Estimating the potential impact of COVID-19-related disruptions on HIV incidence and mortality among men who have sex with men in the United States: a modelling study

**DOI:** 10.1101/2020.10.30.20222893

**Authors:** Kate M Mitchell, Dobromir Dimitrov, Romain Silhol, Lily Geidelberg, Mia Moore, Albert Liu, Chris Beyrer, Kenneth H. Mayer, Stefan Baral, Marie-Claude Boily

**Affiliations:** MRC Centre for Global Infectious Disease Analysis, School of Public Health, Imperial College London, London, United Kingdom; HIV Prevention Trials Network Modelling Centre, Imperial College London, London, United Kingdom; Vaccine and Infectious Disease Division, Fred Hutchinson Cancer Research Center, Seattle, Washington, United States; Department of Medicine, University of California San Francisco, San Francisco, California, United States; Bridge HIV, Population Health Division, San Francisco Department of Public Health, San Francisco, California, United States; Department of Epidemiology, Johns Hopkins Bloomberg School of Public Health, Baltimore, Maryland, United States; The Fenway Institute, Fenway Health, Boston, Massachusetts, United States; Harvard Medical School and T.C. School of Public Health, Boston, Massachusetts, United States

## Abstract

**Background:** During the COVID-19 pandemic, gay and other men who have sex with men (MSM) in the United States (US) report similar or fewer sexual partners and reduced HIV testing and care access. Pre-exposure prophylaxis (PrEP) use has declined. We estimated the potential impact of COVID-19 on HIV incidence and mortality among US MSM.

**Methods:** We used a calibrated HIV transmission model for MSM in Baltimore, Maryland, and available data on COVID-19-related disruptions to predict impacts of data-driven reductions in sexual partners(0%,25%,50%), condom use(5%), HIV testing(20%), viral suppression(10%), PrEP initiations(72%), PrEP use(9%) and ART initiations(50%), exploring different disruption durations and magnitudes. We estimated the median (95% credible interval) change in cumulative new HIV infections and deaths among MSM over one and five years, compared with a scenario without COVID-19-related disruptions.

**Findings:** A six-month 25% reduction in sexual partners among Baltimore MSM, without HIV service changes, could reduce new HIV infections by 12·2%(11·7,12·8%) and 3·0%(2·6,3·4%) over one and five years, respectively. In the absence of changes in sexual behaviour, the six-month data-driven disruptions to condom use, testing, viral suppression, PrEP initiations, PrEP use and ART initiations combined were predicted to increase new HIV infections by 10·5%(5·8,16·5%) over one year, and by 3·5%(2·1,5·4%) over five years. A 25% reduction in partnerships offsets the negative impact of these combined service disruptions on new HIV infections (overall reduction 3·9%(−1·0,7·4%), 0·0%(−1·4,0·9%) over one, five years, respectively), but not on HIV deaths (corresponding increases 11·0%(6·2,17·7%), 2·6%(1·5,4·3%)). The predicted impacts of reductions in partnerships or viral suppression doubled if they lasted 12 months or if disruptions were twice as large.

**Interpretation:** Maintaining access to ART and adherence support is of the utmost importance to minimise excess HIV-related mortality due to COVID-19 restrictions in the US, even if accompanied by reductions in sexual partnerships.

**Funding:** NIH

**Research in context:** *Evidence before this study:* The COVID-19 pandemic and responses to it have disrupted HIV prevention and treatment services and led to changes in sexual risk behaviour in the United States, but the overall potential impact on HIV transmission and HIV-related mortality is not known. We searched PubMed for articles documenting COVID-related disruptions to HIV prevention and treatment and changes in sexual risk behaviour in the United States, published between 1^st^ January and 7^th^ October 2020, with no language restrictions, using the terms COVID* AND (HIV OR AIDS) AND (“United States” OR US). We identified three cross-sectional surveys assessing changes in sexual risk behaviour among men who have sex with men (MSM) in the United States, one finding a reduction, one a slight increase, and one no change in partner numbers during COVID-19 restrictions. Two of these studies also found reductions in reported HIV testing, HIV care and/or access to pre-exposure prophylaxis (PrEP) among MSM due to COVID-19. A separate study from a San Francisco clinic found declines in viral suppression among its clients during lockdown. We searched PubMed for articles estimating the impact of COVID-related disruptions on HIV transmission and mortality published between 1^st^ January 2020 and 12^th^ October 2020, with no language restrictions, using the following terms: COVID* AND model* AND (HIV OR AIDS). We identified two published studies which had used mathematical modelling to estimate the impact of hypothetical COVID-19-related disruptions to HIV programmes on HIV-related deaths and/or new HIV infections in Africa, another published study using modelling to estimate the impact of COVID-19-related disruptions and linked HIV and SARS-CoV-2 testing on new HIV infections in six cities in the United States, and a pre-print reporting modelling of the impact of COVID-19-related disruptions on HIV incidence among men who have sex with men in Atlanta, United States. None of these studies were informed by data on the size of these disruptions. The two African studies and the Atlanta study assessed the impact of disruptions to different healthcare disruptions separately, and all found that the greatest negative impacts on new HIV infections and/or deaths would arise from interruptions to antiretroviral therapy. They all found smaller effects on HIV-related mortality and/or incidence from other healthcare disruptions, including HIV testing, PrEP use and condom supplies. The United States study assessing the impact of linked HIV and SARS-CoV-2 testing estimated that this could substantially reduce HIV incidence.

*Added value of this study:* We used mathematical modelling to derive estimates of the potential impact of the COVID-19 pandemic and associated restrictions on HIV incidence and mortality among MSM in the United States, directly informed by data from the United States on disruptions to HIV testing, antiretroviral therapy and pre-exposure prophylaxis services and reported changes in sexual risk behaviour during the COVID-19 pandemic. We also assessed the impact of an HIV testing campaign during COVID-19 lockdown.

*Implications of all the available evidence:* In the United States, maintaining access to antiretroviral therapy and adherence support for both existing and new users will be crucial to minimize excess HIV-related deaths arising from the COVID-19 pandemic among men who have sex with men. While reductions in sexual risk behaviour may offset increases in new HIV infections arising from disruptions to HIV prevention and treatment services, this will not offset the additional HIV-related deaths which are also predicted to occur. There are mixed findings on the impact of an HIV testing campaign among US MSM during COVID-19 lockdown. Together, these studies highlight the importance of maintaining effective HIV treatment provision during the COVID-19 pandemic.

## Introduction

The COVID-19 pandemic and responses to it have disrupted HIV prevention and treatment services around the world, and influenced sexual behaviour, with consequences for HIV transmission and mortality.^1-4^

Mathematical modelling can draw together information on HIV epidemiology and care, and disruptions caused by COVID-19, to estimate the potential impact of these disruptions on HIV transmission and mortality, and identify where efforts should be prioritised.^5^ Modelling for Africa suggests a six-month interruption of antiretroviral therapy (ART) drugs due to COVID-19 across 50% of people living with HIV could lead to 39-87% more HIV-related deaths over the next year.^6^ Modelling studies for the United States (US) projected increases in HIV incidence due to disruptions to HIV screening, pre-exposure prophylaxis (PrEP) and ART could be offset by reductions in sexual activity of similar magnitude and duration.^7,8^

In the US, COVID-19 prevention and mitigation programs have included stay-at-home orders and venue closures that may limit access to in-person medical care and opportunities to meet sexual partners in other households.^9,10^ Reduced provision of in-person medical care has been reported as healthcare staff were diverted to the COVID-19 response and physical distancing implemented,^11-14^ and decreased viral suppression seen among people living with HIV in San Francisco.^15^ In Baltimore, Maryland, which has very high HIV prevalence among gay, bisexual and other men who have sex with men (MSM; 37% in 2017^16^), stay-at-home orders were in effect from March 30^th^ to June 8^th^ 2020. In late April, reduced HIV and STI testing were reported in Baltimore, with many health department staff focussing on the COVID-19 response.^11^ MSM in the US – among whom 70% of new HIV infections occur nationally – reported fewer sexual partners because of COVID-19 in two national surveys^2,10^, but slightly increased or stable partner numbers in two other surveys,^17,18^ all conducted in April-May 2020. In these surveys, MSM also reported reduced access to HIV testing,^2,17^ care^2^ and PrEP^2,17^ due to COVID-19. Although HIV and bacterial STI screening at a Boston community health centre decreased by 81% early in the pandemic, gonorrhoea and chlamydia test positivity rate increased, suggesting ongoing risk-taking behaviour by a subset of the population.^12^

In the United Kingdom, an HIV testing campaign was launched encouraging MSM and other groups at risk of HIV acquisition to test for HIV during lockdown using home-testing kits, to take advantage of temporary reductions in sexual risk behaviour and hoping to break chains of HIV transmission.^19^ Modelling for the US suggests that offering HIV testing alongside SARS-CoV-2 testing could reduce HIV incidence.^8^

We used mathematical modelling together with MSM-specific survey and clinic data to estimate the potential impact of the COVID-19 pandemic and responses to it on new HIV infections and HIV-related deaths among MSM in Baltimore, and to identify where HIV prevention and treatment efforts should be focussed during the COVID-19 epidemic to most effectively mitigate negative impacts of COVID-19 on HIV transmission and survival for this population. We assessed the extent to which reductions in sexual risk behaviour might offset the impacts of reduced access to HIV care and prevention. We also estimated the potential impact of a hypothetical HIV testing campaign during lockdown in this setting.

## Methods

### Model

We adapted a previously-published deterministic, compartmental model of sexual HIV transmission and treatment among US MSM,^20^ to include PrEP use. Briefly, the modelled population is divided into mutually exclusive compartments, stratified by age, race, PrEP use, HIV infection stage, set-point viral load, and HIV care engagement. In the model, HIV transmission occurs through main, casual and commercial sexual partnerships, as a function of numbers of sexual partners, sex acts per partnership, condom and PrEP use, and HIV infection and ART status of sexual partners. Model equations, schematics, and detailed descriptions are given in the Supplementary Material.

### Model calibration

The model has previously been calibrated to data for MSM in Baltimore.^20^ Briefly, the model was parameterised with demographic, sexual risk behaviour and HIV testing data from National HIV Behavioral Surveillance (NHBS) data for MSM in Baltimore (2004-2014), and fitted to demographic, HIV prevalence and ART coverage data from Baltimore MSM NHBS and Maryland Department of Health data on HIV care access and viral suppression for Baltimore MSM, using a Bayesian approach. From this we obtained 169 unique parameter combinations (‘fits’) giving outputs consistent with HIV prevalence, demography and ART coverage/viral suppression data trends. For the current analysis, we parameterised age- and race-specific PrEP adherence and dropout using US PrEP Demo Project data^21^ and calibrated the existing 169 model fits to NHBS PrEP coverage estimates, by adjusting PrEP uptake rates. Key parameters and care continuum levels in the model fits are given in Table 1. See Supplementary Material for full parameter and fitting data tables and plots of model fits.

**Table 1.** Summary of key sexual behaviour and care continuum parameters and care continuum levels in model fits – without COVID-19-related disruptions.

| Parameters – fitted values (2020 level) | Range |
| --- | --- |
| **Sexual behaviour parameters** |  |
| Average number of new main anal sex partners per year |  |
| 18-24-year-old black MSM | 0.58-0.8 |
| >24-year-old black MSM | 0.36-0.57 |
| 18-24-year-old white MSM | 0.08-0.37 |
| >24-year-old white MSM | 0.11-0.21 |
| Average number of new casual anal sex partners per year |  |
| 18-24-year-old black MSM | 1.54-2.08 |
| >24-year-old black MSM | 0.81-1.24 |
| 18-24-year-old white MSM | 0.05-0.93 |
| >24-year-old white MSM | 0.29-1.07 |
| Average number of new commercial anal sex partners per year |  |
| 18-24-year-old black MSM | 0-1.35 |
| >24-year-old black MSM | 0.15-0.85 |
| 18-24-year-old white MSM | 0-0.28 |
| >24-year-old white MSM | 0-0.07 |
| Percentage of sex acts in which condom used |  |
| Main partnerships, both partners black | 47-67 |
| Main partnerships, either partner white | 30-39 |
| Casual partnerships (any race partner) | 63-72 |
| Commercial partnerships (any race partner) | 21-78 |
| **Care continuum parameters** |  |
| Percentage of undiagnosed MSM (not on PrEP) testing for HIV per year |  |
| 18-24-year-old black MSM | 25.6-94.8 |
| >24-year-old black MSM | 20.4-70.1 |
| 18-24-year-old white MSM | 14.2-81.7 |
| >24-year-old white MSM | 13.8-69.5 |
| **Care continuum levels in model fits (2020 level)** |  |
| Percentage of HIV-positive MSM diagnosed | 74-95 |
| Percentage of diagnosed MSM on ART | 43-87 |
| Percentage of MSM on ART who are virally suppressed | 76-93 |
| Percentage of HIV-negative MSM on PrEP | 10-21 |

### Scenarios

#### Base case scenario

The base case scenario – to which COVID-19 disruption scenarios are compared – represented the expected course of the HIV epidemic over time if the COVID-19 pandemic and related disruptions had not occurred. Rates of HIV testing, ART and PrEP initiation, adherence and dropout, sexual partner change and condom use were all assumed to be maintained at their 2019 level from 2020 onwards. Levels of viral suppression and PrEP use increase over 2020-2025 in the base case scenario (Figure S4) due to declines in HIV incidence and earlier increases in PrEP initiation rates.

#### Disruptions due to COVID-19 and responses to COVID-19

We modelled data-driven disruptions to HIV testing, condom use, viral suppression and sexual partnerships due to COVID-19, informed by available data from two online surveys of MSM in the US,^2,10^ conducted while stay-at-home orders were in place (in April/May 2020) and asking about COVID-19-related changes to sexual health, sexual risk behaviour and HIV prevention and care (Table 2). In these two studies, 51%^2^ and 40%^10^ of MSM reported having fewer sexual partners due to COVID-19 (only 1% reported having more partners^2,10^), but the studies did not ask how many fewer partners they had. As those reporting fewer partners may still have sexual partners, we explored overall reductions in numbers of all types of sexual partners of 25% and 50% (equivalent to 50% and 100% reduction among 50% of MSM). As two other online surveys of US MSM found no change in numbers of casual sexual partners^18^ or slight increases in total but no change in unprotected sexual partner numbers,^17^ we also considered scenarios with no change in partner numbers. We modelled observed reductions in PrEP initiations, PrEP use and HIV testing among those on PrEP, based upon a study at a Boston PrEP clinic.^12^ Where possible, we used any age- and/or race-stratified estimates from these studies to inform age- and race-specific estimates for these disruptions (Table 2, Table S3). In addition, we used data from a global online survey of MSM on ART access, stratified by whether MSM were members of a racial/ethnic minority,^22^ to estimate race-specific relative reductions in viral suppression. As none of these studies assessed changes to ART initiations, we assumed these were reduced by 50%, following qualitative reports of HIV care disruptions in Baltimore.^11^

**Table 2:** Summary of magnitudes of disruptions modelled, with sources.

| **Disruption to:** | **Overall data-driven reduction used in main scenario** | **Age- and/or race-specific reductions used in main scenario** | **Overall reduction explored in sensitivity analysis** | **Source** |
| --- | --- | --- | --- | --- |
| HIV testing | 20% | 18-24-year-olds: 25%<br>≥25-year-olds: 19% | 50%, 75%, 100% | Reported access to testing and trouble getting an HIV test, overall and by age, survey of US MSM.<sup>2</sup><br>Consistent with reported prevented access to HIV testing in another survey of US MSM<sup>17</sup> |
| ART initiations | 50% | .. | 50%, 75%, 100% | Assumption |
| Viral suppression | 10% | White: 9%<br>Black: 15% | 10%, 25%, 50% | Reported % having fewer viral load or other lab tests, reduced access to HIV medications, trouble getting an ART prescription, taking medications daily less often, from survey of US MSM.<sup>2</sup><br>Race differences from reported ART access in a global MSM survey<sup>22</sup> |
| PrEP initiations | 72% | .. | 50%, 75%, 100% | Numbers of PrEP initiations at a Boston PrEP clinic<sup>12</sup> |
| PrEP use | 9% | Black 18-24-yr-olds:13%<br>White 18-24-yr-olds:11%<br>Black ≥25-yr-olds: 9%<br>White ≥25-yr-olds: 8% | 10%, 25%, 50% | Numbers of excess missed PrEP refills during lockdown out of the total number of PrEP patients at a Boston PrEP clinic,<sup>12</sup> reported trouble getting a PrEP prescription or medication from surveys of US MSM.<sup>2,17</sup><br>Age and race differences from % missed PrEP refills by age and race during lockdown at a Boston PrEP clinic.<sup>12</sup> |
| HIV testing on PrEP | 85% | .. | 50%, 75%, 100% | From numbers of HIV tests conducted at a Boston PrEP clinic<sup>12</sup> |
| Condom use | 5% | .. | 10%, 25%, 50% | From reported change in condom use, survey of US MSM<sup>2</sup> |
| Partner numbers | 25%, 50% | 18-24-year olds: 22%, 44%<br>≥25-year-olds: 26%, 52% | 10%, 25%, 50% | From % reporting fewer sex partners in surveys of US MSM,<sup>2,10</sup> assuming these reduced their number of partners by 50-100%. Age differences from % reporting fewer sex partners in survey of US MSM<sup>2</sup> |

These changes were represented in the model by reducing rates of HIV testing, ART and PrEP initiation, and levels of condom use and partner numbers by the estimated amount. Reductions in PrEP use were based upon data on PrEP refills at a Boston PrEP clinic,^12^ and data on access to PrEP prescriptions and medication from online surveys of US MSM,^2,17^ and were represented in the model by reductions in PrEP adherence. Reductions in viral suppression among those on ART were represented by increases in HIV transmissibility and mortality for those in the virally suppressed compartment, weighted by the proportion of people assumed to no longer be virally suppressed (who were assumed to have the same infectiousness and mortality as those not on ART).

In our main analysis, we modelled the impact of a six-month disruption due to COVID-19, assuming all disruptions were reversed at the end of the period, with parameters reset instantaneously to their pre-disruption levels.

We assessed the impact of each disruption separately and in combination.

### Sensitivity analysis

We tested the sensitivity of our findings to the magnitude of the different disruptions, comparing the individual impacts of 10%, 25% and 50% reductions in partner numbers, condom use, PrEP use and viral suppression, and 50%, 75% and 100% reductions in ART and PrEP initiation rates and HIV testing on and off PrEP. We also explored the impact of three- and 12-month disruptions.

### HIV testing campaign

We assessed the expected impact of an HIV testing campaign during lockdown, based on the UK Test Now, Stop HIV campaign,^19^ which aimed to capitalise on temporary declines in sexual activity and HIV transmission to identify those with undiagnosed HIV and prevent further HIV transmission. We modelled this testing campaign alongside plausible COVID-related disruptions to (a) sexual partner numbers only or (b) sexual partner numbers, ART initiation, viral suppression, PrEP initiation, PrEP use and condom use, for a six-month period. We simulated HIV testing campaigns in which 90% of MSM tested at least once during the six-month disruption period, after which HIV testing rates returned to pre-disruption levels.

### Outcome measures

We estimated short-term (one year) and long-term (five year) impacts of COVID-19 on cumulative new HIV infections and HIV-related deaths measured from the start of COVID-related disruptions. Impacts were calculated relative to the base case scenario and expressed as absolute or percentage change.

The transmission model was coded and run in C++ and calculations were conducted using R version 3.6.2.

## Results

### Predicted impact of data-driven six-month disruptions

We first assessed the impact of data-driven six-month COVID-related disruptions on the number of new infections (Figure 1a) and HIV-related deaths (Figure 1b). We found reducing sexual partner numbers by 25% could substantially reduce new HIV infections, by a median 12·2% (95%CrI 11·7, 12·8%) and 3·0% (2·6, 3·4%) over the following one and five years, respectively, than if the COVID-19 pandemic and associated disruptions had not occurred (Figure 1a). Twice that impact was seen for a 50% reduction in partnerships (Figure 1a). Conversely, data-driven 10% reductions in levels of viral suppression among those on ART were predicted to increase new infections the most, by 6·4% (2·6, 11·9%) over one year and 1·5% (0·5, 3·1%) over five years, with some uncertainty (Figure 1a). Explored reductions in ART initiations (by 50%) and other disruptions to HIV services suggested by data including reductions in HIV testing (20%), PrEP initiations (72%), HIV testing on PrEP (85%), condom use (5%), and PrEP use (9%) each had a smaller negative impact, increasing new infections by less than 2% on average over one year (Figure 1a). Predicted relative increases in new HIV infections decreased over five years for all disruptions, with larger reductions in relative impact over time for disruptions to condom use, viral suppression and partner numbers (Figure 1a).

**Figure 1:**
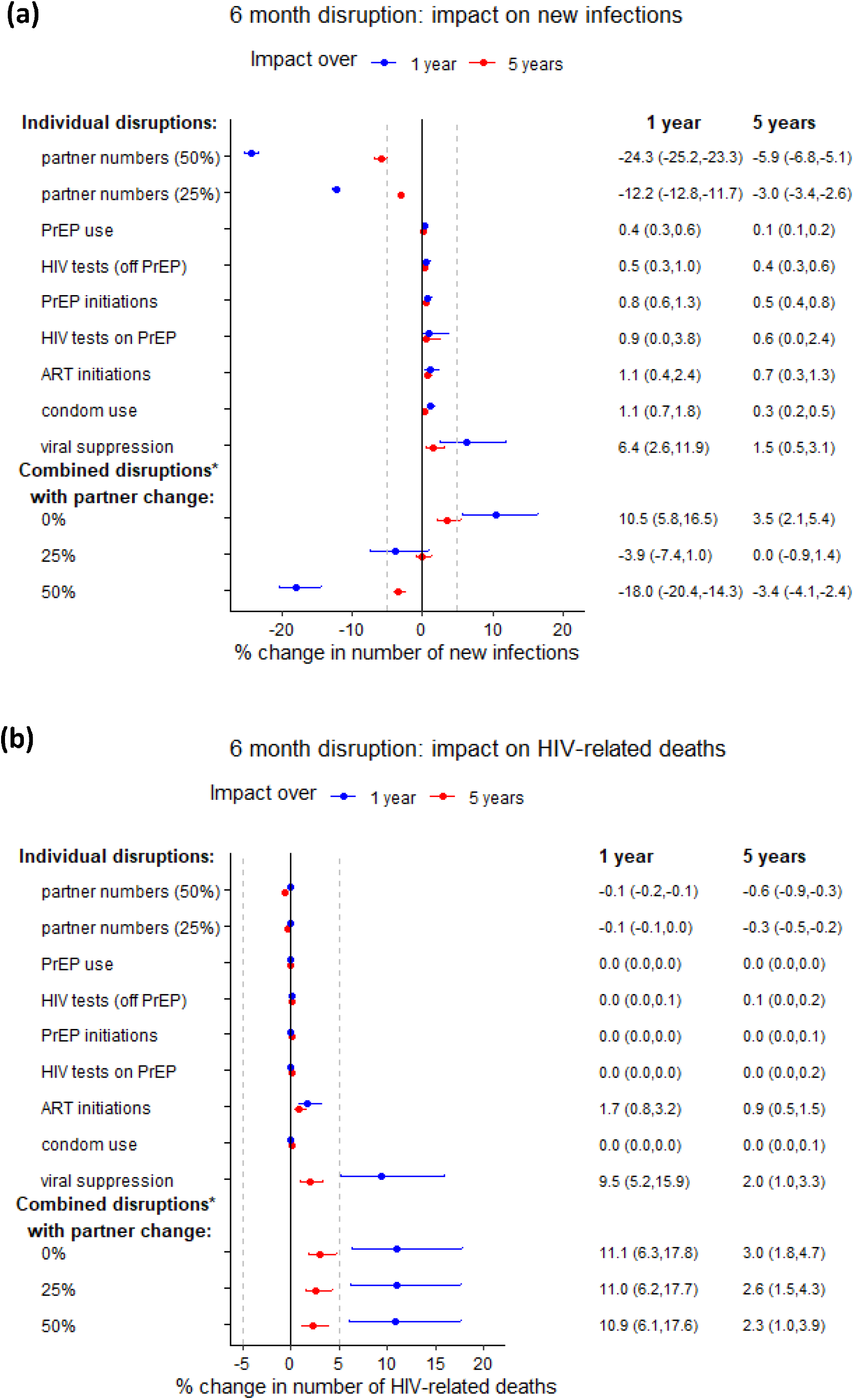
Impact of 6-month individual and combined estimated disruptions due to COVID-19. Estimated disruptions are based on available data (see Table 2, Table S3 for details). Impact on (a) cumulative new HIV infections and (b) cumulative HIV-related deaths, over 1 year (blue points) and 5 years (red points). Points are median and error bars are 95% credible intervals across all model fits. Disruptions are assumed to last for 6 months. Individual overall disruption magnitudes are: 20% reduction in HIV tests, 5% reduction in condom use, 72% reduction in PrEP initiations, 9% reduction in PrEP use, 85% reduction in HIV testing on PrEP, 50% reduction in new ART initiations and 10% reduction in viral suppression (see Table 2 for age- and race-specific disruptions). *Combined disruption consists of 20% reduction in HIV tests, 5% reduction in condom use, 72% reduction in PrEP initiations, 9% reduction in PrEP refills, 85% reduction in HIV testing on PrEP, 50% reduction in new ART initiations and 10% reduction in viral suppression. Dashed vertical lines are at −5% and 5%

Without any change in sexual behaviour, the six-month disruptions to testing, ART initiations, viral suppression, PrEP initiations, PrEP use and condom use combined were predicted to increase new HIV infections by 10·5% (5·8, 16·5%) over one year, and by 3·5% (2·1, 5·4%) over five years. The impact of these combined disruptions could be outweighed by a concurrent six-month 25% reduction in partner numbers (overall median 3·9% decrease in new infections over one year, median 0·0% change over five years; Figure 1a).

We predicted substantial increases in HIV-related deaths from disruptions to ART initiations and especially viral suppression among those on ART, associated with 1·7% (0·8, 3·2%) and 9·5% (5·2, 15·9%) more HIV-related deaths over one year, respectively, with substantial uncertainty (Figure 1b). The other individual data-driven disruptions – to HIV testing, PrEP and condom use, PrEP initiation and partner numbers – would have minimal impact on deaths (<1% change over one or five years). The combined predicted effect of all the disruptions to HIV-related services was predicted an 11·1% (6·3, 17·8%) increase in HIV-related deaths over one year, which equates to 4·8 (2·2-9·0) additional deaths in this population of around 2000 HIV-infected MSM over only one year. The proportional combined impact was almost 4-fold smaller over five years than over one year, with 6·3 (2·8-11·5) additional deaths over five years. Unlike new infections, increases in HIV-related deaths over one or five years were not offset by reductions in sexual risk behaviour (impact reduced by <1 percentage point (pp); Figure 1b). This is because reduced sexual risk behaviour has only an indirect and delayed effect on HIV-related deaths, through reducing new HIV infections (Figure 1b).

### Sensitivity analysis

For individual disruptions, we found each of the following: a 10% reduction in partner numbers, 25% less condom use, or a 10% reduction in viral suppression, could alter the number of new HIV infections over one year by more than 5% (Figure 2a). In contrast, even complete cessation (100% reduction) of HIV testing, ART initiation or PrEP initiation, or a 50% reduction in PrEP use individually would be insufficient to increase new HIV infections by more than 5% on average. Complete cessation of ART initiations or a 10% reduction in viral suppression would result in a 5% or greater increase in HIV-related deaths over one year (Figure 2b).

**Figure 2:**
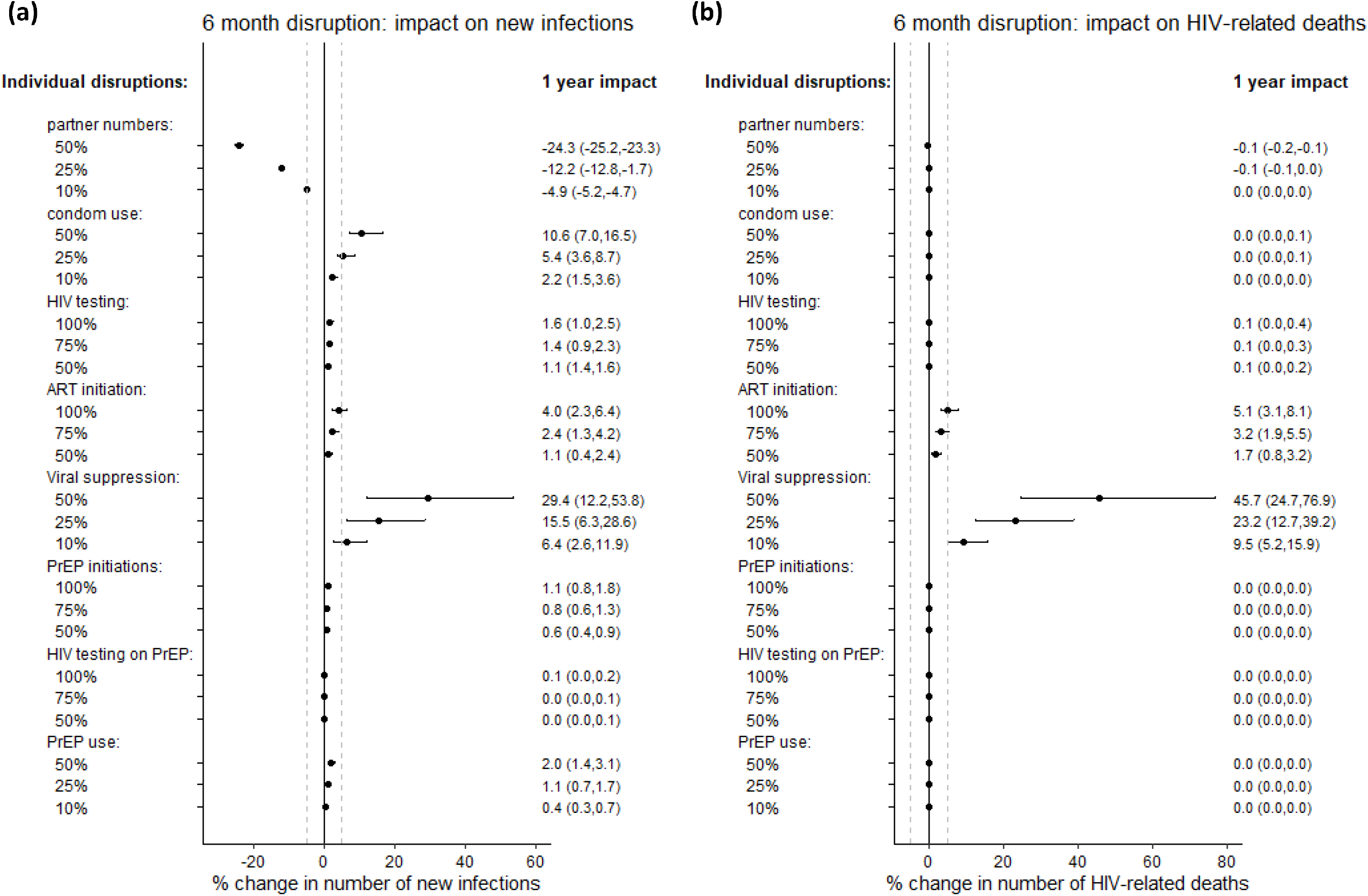
Sensitivity analysis on disruption magnitude: 1-year impact. Impact of individual disruptions due to COVID-19, exploring different values for the size of these disruptions, indicated on plot. Impact on (a) cumulative new HIV infections and (b) cumulative HIV-related deaths, over 1 year. Points are median and error bars are 95% credible intervals across all model fits. Disruptions are assumed to last for 6 months. Dashed vertical lines are at −5% and 5%. 5-year impacts are shown in figure S6

Predicted impacts on infections and deaths were proportional to the magnitude of reduction in sexual partnerships, condom use, viral suppression, and PrEP initiation (Figure S6a-d, Figure S7a-d). Impacts of disruptions to ART initiations and HIV testing on PrEP were more than proportional (increasing more with larger disruptions) while the impacts of reduced PrEP use and other HIV testing were less than proportional to disruption magnitude (Figure S6e-h,Figure S7e-h).

The predicted impact of disruptions affecting partner numbers, condom use, viral suppression and PrEP use on new HIV infections also varied proportionately with disruption duration, measured over both one and five years (Figure 3a-d). Impacts of disruptions to PrEP and ART initiation and HIV testing varied proportionately with disruption duration when measured over five years, but not over one year, when impacts were less than proportional to disruption duration (Figure 3d-h). Similar patterns were seen for HIV-related deaths (Figure S8).

**Figure 3:**
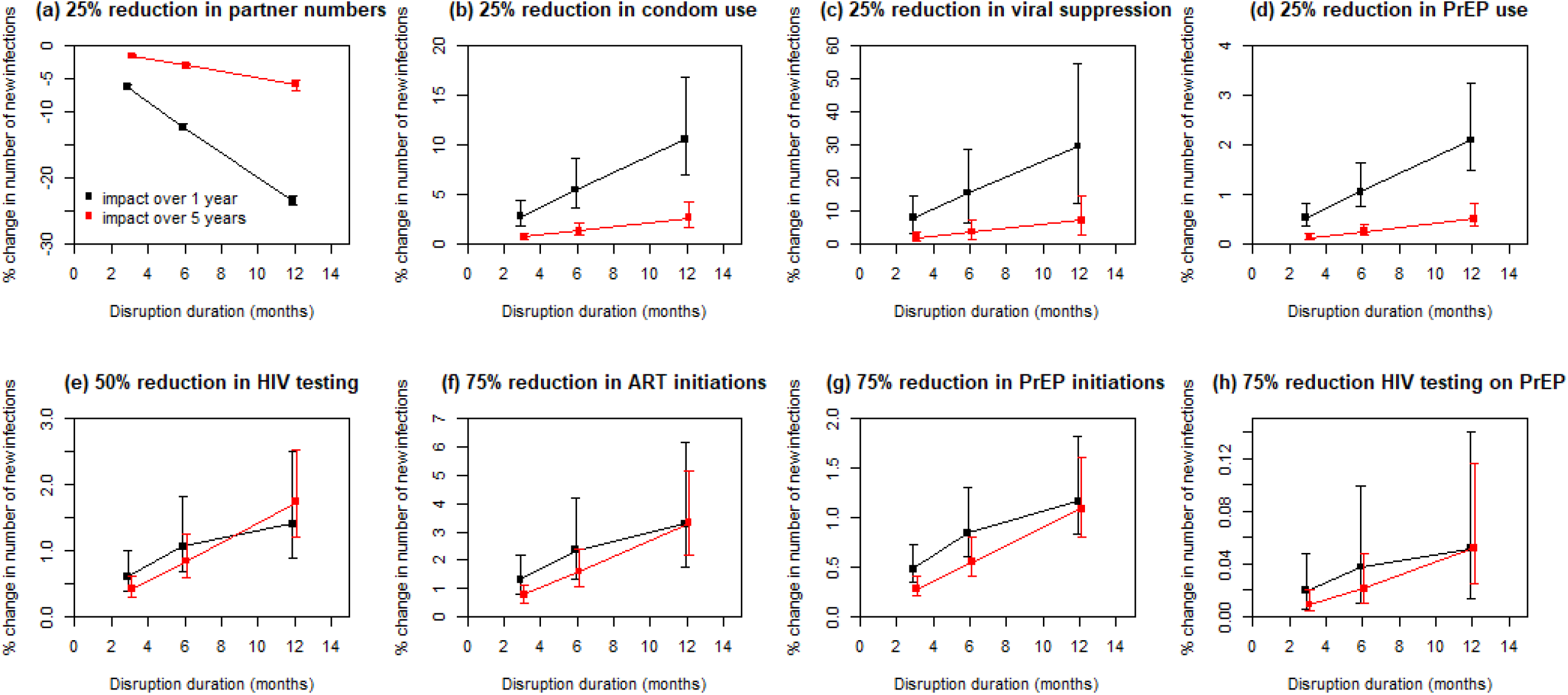
Sensitivity analysis on disruption duration. Impact on cumulative new HIV infections over 1 year (black points) and 5 years (red points), for disruptions indicated, over 3, 6 and 12 months. Points are median and error bars are 95% credible intervals across all model fits. Note very different y-axis scales used to clearly show linearity of trends.

### Impact of an HIV testing campaign

If COVID-19 restrictions only reduced sexual activity, by 25%, without impacting HIV care and prevention services, then successfully testing 90% of MSM at least once during the six-month COVID-related disruption may avert 3·7pp (2·3, 13·4pp) and 2.9pp (1·7, 14·7pp) additional new infections over one or five years in addition to the infections averted by reduced partner numbers (Figure 4a,b; Figure S9a,b). However, taking into account all other likely COVID-19 disruptions alongside a 25% reduction in sexual activity, such an HIV testing campaign is expected to have a very modest impact, averting only 0·9pp (0·0, 6·4pp) and 0·8pp (−0·1, 9·3pp) more infections (compared with those averted without the testing campaign) over one or five years (Figure 4c,d; Figure S9c,d). The testing campaign was predicted to have a greater impact (2-10pp more infections averted over five years) for scenarios with lower pre-COVID levels of awareness of HIV-positive status (<85%; Figure S10).

**Figure 4:**
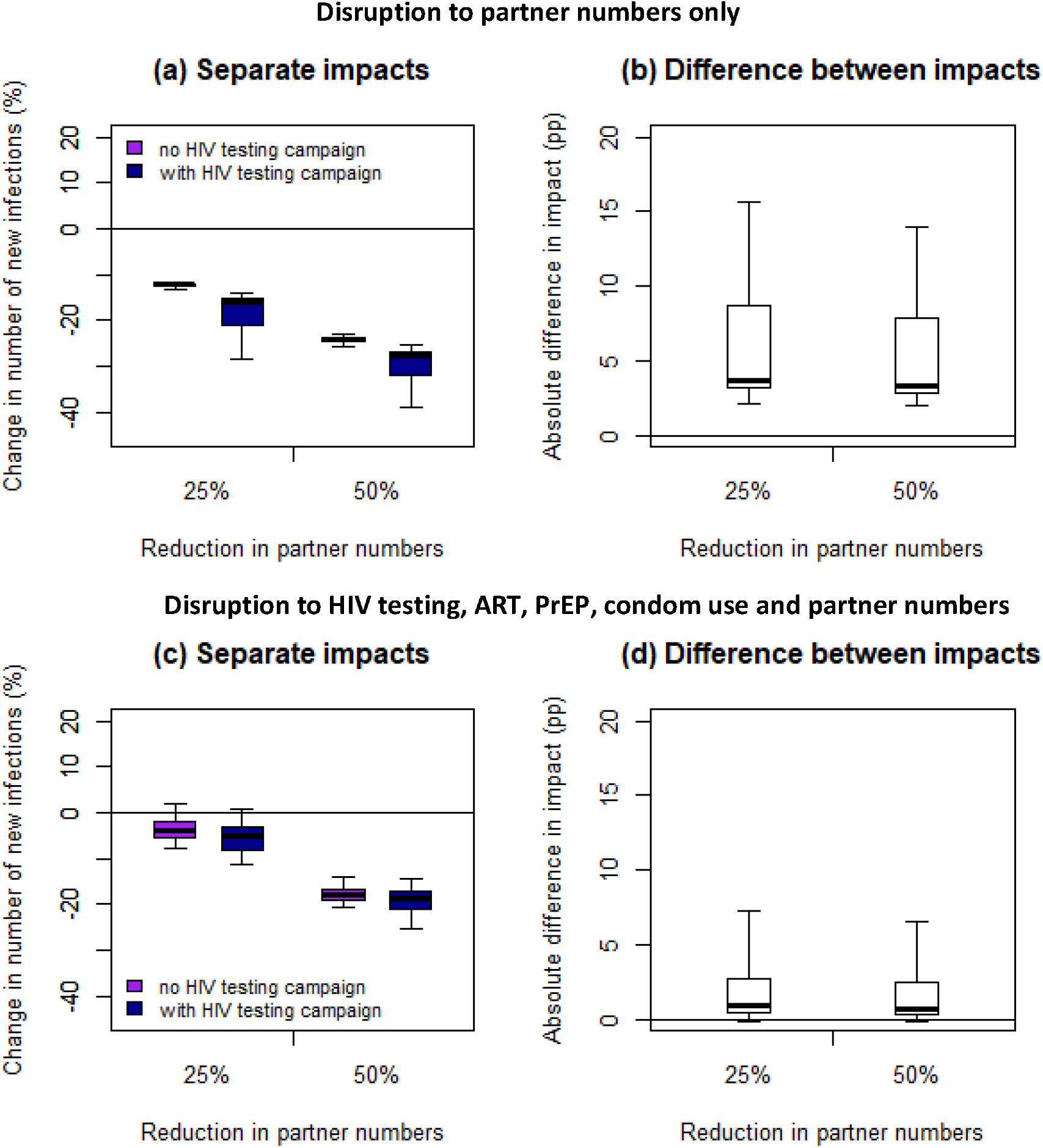
Impact of disruptions with and without additional HIV testing campaign, over 1 year. Impact on cumulative new HIV infections over one year of six-month disruptions to (a,b) partner numbers only or (c,d) HIV testing, ART initiation, viral suppression, PrEP initiation and continuation and condom use as well as partner numbers, with (dark blue bars) or without (purple bars) an additional HIV testing campaign reaching 90% of MSM for HIV testing during the six-month disruption. Panels (a) and (c) show the impacts relative to the base case scenario, with no COVID, and panels (b) and (d) show the difference in percentage points (pp) between the impacts of the disruptions with and without the additional HIV testing campaign. For the full disruption (c,d), we modelled a 20% reduction in HIV tests (in the absence of the HIV testing campaign), 5% reduction in condom use, 72% reduction in PrEP initiations, 40% reduction in PrEP refills, 85% reduction in HIV testing on PrEP, 100% reduction in new ART initiations and 10% reduction in viral suppression (see Table 2 for age- and race-specific disruptions). Thick lines are median, boxes are interquartile range, and whiskers full range across all model fits.

## Discussion

Our modelling results predict that modest reported COVID-19-related disruptions to HIV testing, PrEP use, condom use and viral suppression, and larger disruptions to PrEP and ART initiations, without any change in sexual risk behaviour (as suggested by some surveys), could lead to substantial short-term increases (11% over one year for a six-month disruption) in new HIV infections and HIV-related deaths among Baltimore MSM. Our results suggest a 25% or 50% reduction in sexual partner numbers (consistent with other survey data) could offset increases in new HIV infections arising from COVID-19-related disruptions to HIV services, but would not offset additional HIV-related deaths caused by these service disruptions. Together, these six-month service disruptions alongside a 25% reduction in partner numbers are predicted to give little change in new HIV infections (−4% over one year and 0% over five years). However, even with a 50% reduction in partner numbers, these same disruptions may still give rise to substantial short-term increases in HIV-related deaths (11%, equivalent to five additional deaths, over one year).

Our sensitivity analysis suggests disruptions to viral suppression and condom use would have the greatest adverse effects on new HIV infections, while reductions in viral suppression and new ART initiations could lead to the most HIV-related deaths. Even a 10% reduction in viral suppression among those on ART, lasting six months, would lead to ≥5% increases in both new HIV infections and deaths over the following year. Condom use would need to be reduced by 25% to cause a 5% increase in new HIV infections. ART initiations would need to cease completely for six months to bring about a 5% increase in annual HIV-related deaths. Increasing the duration of disruptions to viral suppression and condom use from six to 12 months could double their impact on both HIV infections and deaths, as could doubling the size of each disruption. Finally, we find a widespread HIV testing campaign during lockdowns in this setting is likely to have only a small incremental impact on HIV incidence, due to concurrent disruptions to ART, PrEP and condom use.

Our results suggest that, during COVID-19-related restrictions, HIV prevention and treatment efforts should be focussed upon maintaining access to ART and ART adherence support for both new and existing users, to minimize negative impacts of COVID-19 on new HIV infections and, especially, HIV-related mortality. Multi-month ART dispensing and tele-health services may be sufficient to help ART users who are already stably virally suppressed maintain suppression during lockdown. Triaging to prioritise in-person care and viral load tests to new or less stably-suppressed ART patients will also be needed,^13,23^ particularly for homeless individuals.^15^ Although US MSM report only minimal reductions in condom use due to COVID-19,^2^ our sensitivity analysis suggests maintaining condom use will be important to prevent extra HIV transmission.

There is uncertainty about the magnitude and durations of disruptions to HIV prevention and treatment services, and how these may change during the COVID-19 pandemic. Going forwards, service provision could improve as services adapt.^12,23^ Conversely, HIV service access could be hampered by loss of employer-provided health insurance as COVID-19 leads to increased unemployment.^2^ Loss of health insurance may be particularly detrimental in states which have not passed the Affordable Care Act. However, ART should be available through the Ryan White Care Program or AIDS Drug Assistance Programs.

There is also uncertainty about how long any reductions in sexual risk behaviour may last. A study among Southern PrEP-using MSM found sexual risk-taking increasing in June 2020 following earlier reductions.^24^ Modelling for Atlanta MSM suggests substantial increases in HIV incidence will occur if service disruptions last longer than sexual behaviour changes.^7^

We find smaller but more persistent impacts (still sizeable after five years) following disruptions to rates of HIV testing, ART and PrEP initiation compared with disruptions to partner numbers and viral suppression. Changes to HIV testing or ART/PrEP initiations rates lead to smaller but longer-lasting changes in diagnosis and ART/PrEP coverage levels, while changes to partner numbers or viral suppression lead to larger direct changes, which we assume rapidly reverse after the disruption period. Post-disruption catch-up campaigns could reduce the longer-term impacts of COVID-19-related disruptions to HIV testing, ART and PrEP initiations.

Several modelling studies have estimated the impact of COVID-19-related disruptions on HIV in sub-Saharan Africa.^6,25-27^ In agreement with our results, these studies all suggest the largest detrimental impact will be due to ART disruptions (equivalent to our viral suppression disruptions), with disruptions to condom use, HIV testing and ART initiations having smaller impacts.^6,25-27^ In an analysis using five different African models, a six-month interruption to ART for 50% of the population was predicted to give a median 9% (range 6-126%) more new HIV infections and 63% (39-87%) more HIV-related deaths over the following year.^6^ Our US estimates for a 50% reduction in viral suppression (29% more infections, 46% more deaths) fall within these ranges. A recent modelling study estimated the impact of COVID-19-related disruptions on HIV incidence among MSM in Atlanta.^7^ Like us, they found reductions in sexual partnerships could offset the impact of service disruptions on new HIV infections, and PrEP reductions had less effect than ART reductions, partly due to low PrEP coverage (15% in Atlanta). ^7^ Another modelling study assessing impacts on HIV incidence in six US cities including Baltimore similarly found that reductions in sexual partnerships could offset health service reductions.^8^ However, in contrast to our findings, they projected substantial reductions in HIV incidence would result from expanded HIV testing in Baltimore during COVID-19-related disruptions – this larger impact may arise from testing among heterosexual populations, who have lower HIV testing rates than MSM, and/or because lower levels of awareness of HIV-positive status were assumed (e.g. 78% among MSM vs. 74-95% in our study).^8^

While our analysis maximised the use of currently available data on the impacts of COVID-19 on HIV prevention and care and sexual risk behaviour for US MSM, including age- and race-specific data, it has some limitations, chiefly due to uncertainty about the exact magnitude and duration of these disruptions. Many of the data informing disruption sizes were collected at a single time-point, and do not capture changes as the COVID-19 pandemic progressed, as discussed above. Some estimates were only semi-quantitative, for example, 50% of MSM reporting fewer partners does not tell us how many fewer partners they had overall. We found no data on reductions in ART initiations. However, our sensitivity analysis explored the impact of different disruption levels and durations. Our HIV transmission model was calibrated to Baltimore, but estimates of disruption magnitude were based on national surveys^2,10^ and a Boston study;^12^ disruptions may differ between states due to different levels of COVID-19, COVID-19 response measures, and health funding. Therefore, we may not have captured the true impact of COVID-19 among Baltimore MSM. Our impact estimates may not be directly applicable to other US locations with different HIV epidemics and services although we expect qualitative insights from our model to still be useful. Our predicted small impact of an HIV testing campaign during lockdown may partly be due to high HIV testing rates already occurring in many of our model fits (based on self-reported testing by Baltimore MSM); we predict larger impacts for populations with lower HIV testing rates. Our small predicted impacts of disruptions to PrEP use may be partly due to low PrEP use among Baltimore MSM (12% in 2017) – larger impacts may be seen in places with greater PrEP use, such as Boston, Chicago, or San Francisco.^28^ We assume reductions in PrEP use and partner numbers occur independently, whereas MSM may be ceasing PrEP use while abstaining from sex or those apparently lapsing may be switching to non-daily use.^29^ In this case, our analysis may overestimate negative impacts of PrEP disruptions. Finally, we do not account for mortality directly caused by COVID-19; we may underestimate mortality impacts as COVID-19 outcomes are worse among patients with co-morbidities or low CD4 counts.^30^

In conclusion, maintaining access to ART and adherence support for those on treatment and those newly diagnosed is crucial to minimise excess HIV-related mortality due to COVID-19-related restrictions in the US. Reductions in viral suppression of 10% could have negative consequences for both HIV incidence and mortality and any declines in sexual risk behaviour would only offset incidence, but not mortality increases. It will be important to collect quantitative data on COVID-19-related effects from multiple time-points and settings, to improve modelling estimates and inform public health decisions. Modelling assessing the impacts of post-COVID ‘catch-up’ campaigns is needed to inform healthcare priorities as restrictions are lifted.

## Data Availability

The data used are given in the supplementary material. The model code is available from the authors on request.

## Author Contributions

KMM, DD, RS, LG and MCB contributed to the conception and design of the modelling analyses. KMM, RS, AL, CB, KHM and SB contributed to identifying and interpreting data sources to inform the disruption effects. KMM conducted the modelling analysis. RS, LG and MCB provided advice on technical aspects of modelling analysis. All authors contributed to interpreting the modelling results. KMM wrote the first draft of the manuscript. All authors contributed to manuscript revision and critical review.

## Acknowledgements

KMM, DD, MM and MCB are supported by the HPTN Modelling Centre, which is funded by the US National Institutes of Health (NIH) [grant number UM1 AI068617] through the HPTN Statistical and Data Management Center. KMM, RS, LG and MCB acknowledge funding from the MRC Centre for Global Infectious Disease Analysis [grant number MR/R015600/1], which is jointly funded by the UK Medical Research Council (MRC) and the UK Foreign, Commonwealth & Development Office (FCDO), under the MRC/FCDO Concordat agreement and is also part of the EDCTP2 programme supported by the European Union. LG is funded by an MRC-DTP scholarship. The funders had no role in the study design, collection, analysis or interpretation of data, writing of the report or the decision to submit the paper for publication.

