## Supplementary Information for "Estimating the potential impact of COVID-19-related disruptions on HIV incidence and mortality among men who have sex with men in the United States: a modelling study"

**SUPPLEMENTARY METHODS**

This description of the methods is adapted from Mitchell et al 2019^1^. The model presented here differs from that presented in Mitchell et al 2019 in also including compartments for MSM on PrEP. Treatment compartments are renumbered here to allow for this inclusion. A brief description of how the model was calibrated to PrEP coverage has been added here, and parameters relating to PrEP efficacy, uptake, adherence and dropout have been added to Table S1. Data used to calibrate the model to PrEP coverage have been added to Table S2, and the model calibration to PrEP coverage has been added to Figure S4.

**Model description**

In the model equations and schematics, uninfected MSM are denoted by $X_{v,w}^{z}$, those with acute HIV infection by $A_{v,w}^{z}$and chronic HIV infection by $Y_{v,w}^{x,y,z}$. Subscripts refer to the following states: $v$ is age group (0 = 18-24 years old; 1 = >24 years old), $w$ is race (0 = black; 1 = white). The younger age group had a lower age limit of 18 to match the minimum age of MSM included in NHBS surveys, which supplied the behavioural parameters and HIV prevalence estimates used in this analysis. Superscripts refer to the following states: $x$ is CD4 count (current CD4 count for those not taking or not adherent to ART, CD4 count at ART initiation for those taking and adherent to ART; 0 = acute, 1 = CD4>500, 2 = CD4 350-500, 3 = CD4 200-350, 4 = CD4 <200 cells/µl), $y$ is set-point viral load (SPVL; 0 = acute, 1 = Log_10_ SPVL<4.0, 2 = Log_10_ SPVL 4.0-4.5, 3 = Log_10_ SPVL 4.5-5.0, 4 = Log_10_ SPVL >5.0), $z$ is care state (0 = never testing, 1 = testing but not diagnosed, 2 = on PrEP, 3 = diagnosed not linked to care, 4 = linked into HIV care, 5 = on ART, adherent and partially suppressed, 6 = in first year on ART, adherent and fully suppressed, 7 = 2^nd^ year on ART adherent and fully suppressed, 8 = 3^rd^ and subsequent years on ART adherent and fully suppressed, 9 = on ART but non-adherent and not suppressed, 10 = stopped taking ART (due to dropout or failure)). For those uninfected with HIV, the only possible care states are $z$=0, 1 or 2. Those with acute infection may be in one of care states $z$=0-5; after achieving full viral suppression on ART they are assumed to no longer be in the acute stage.

Fig S1 shows the age and race groups, with movement and sexual mixing between them. Of individuals entering the sexually active Baltimore MSM population, a proportion $m_{v,w}$ are assumed to be in each combination of age and race group ($m_{v,w}$ is calculated from $m_{black}$, the proportion of incoming MSM who are black, and $m_{young,w}$, the proportion of incoming MSM of each race who are aged 18-24 years old. Those in the 18-24 year old group move into the older age group at an annual rate $\pi_{w}$ per year, corresponding to an average of 1/ $\pi_{w}$ years that sexually active MSM in race group $w$spend in the 18-24 year old age group.

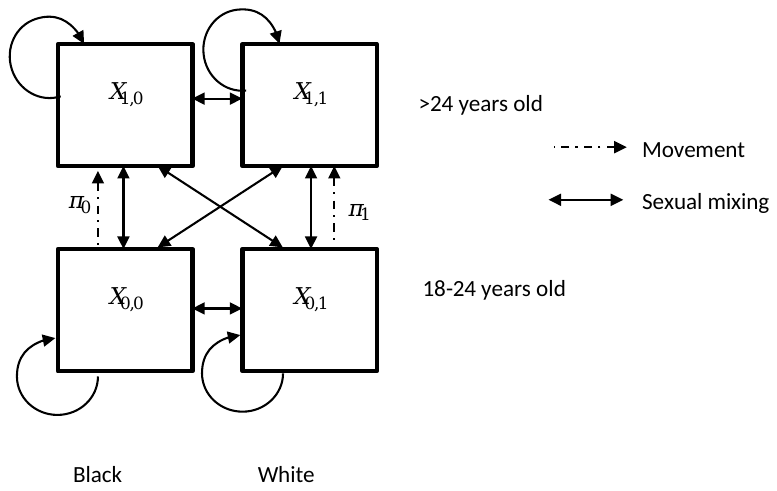

**Fig S1: Age groups, race groups, movement and mixing in the model**

Fig S2 shows the transitions between different stages of HIV infection for those not currently taking ART, by current HIV stage and SPVL. These transitions are the same for all age, race and care states (apart from those on ART and adherent), with the following exceptions: infection rates ($\lambda$) and background death rate ($\mu$) differ by age and race, and infection rates ($\lambda$) also differ by PrEP status.

Susceptible individuals ($X^{z}$) become infected with HIV at a rate $\lambda_{v,w,z}$ and move into the acutely infected compartment ($A^{z}$). After a period (1/$\gamma_{a}$ years) in the acute stage, individuals move into one of 16 compartments ($Y^{x,y,z}$), defined by their SPVL and initial CD4 count after acute infection. A proportion ($\theta_{y}$) of those leaving the acute stage move into SPVL stratum $y$. For each SPVL stratum, a proportion $f_{x,y}$ of those entering SPVL stratum $y$ are initially in CD4 compartment $Y^{x,y,z}$. Within each SPVL stratum, HIV-positive people pass sequentially through progressively lower CD4 count categories. The rate of moving from one CD4 compartment to the next is given by $\gamma_{x,y}$.

There is a constant background per-capita rate of non-HIV related death ($\mu_{v,w}$) from every compartment (susceptibles and all infected compartments), and an additional rate of HIV–related death from each infected compartment ($\alpha_{x,y,z}$), which varies by SPVL and current CD4 count, but takes the same value for all those off ART or non-adherent to ART (z = 0,1,2,3,4,9,10).

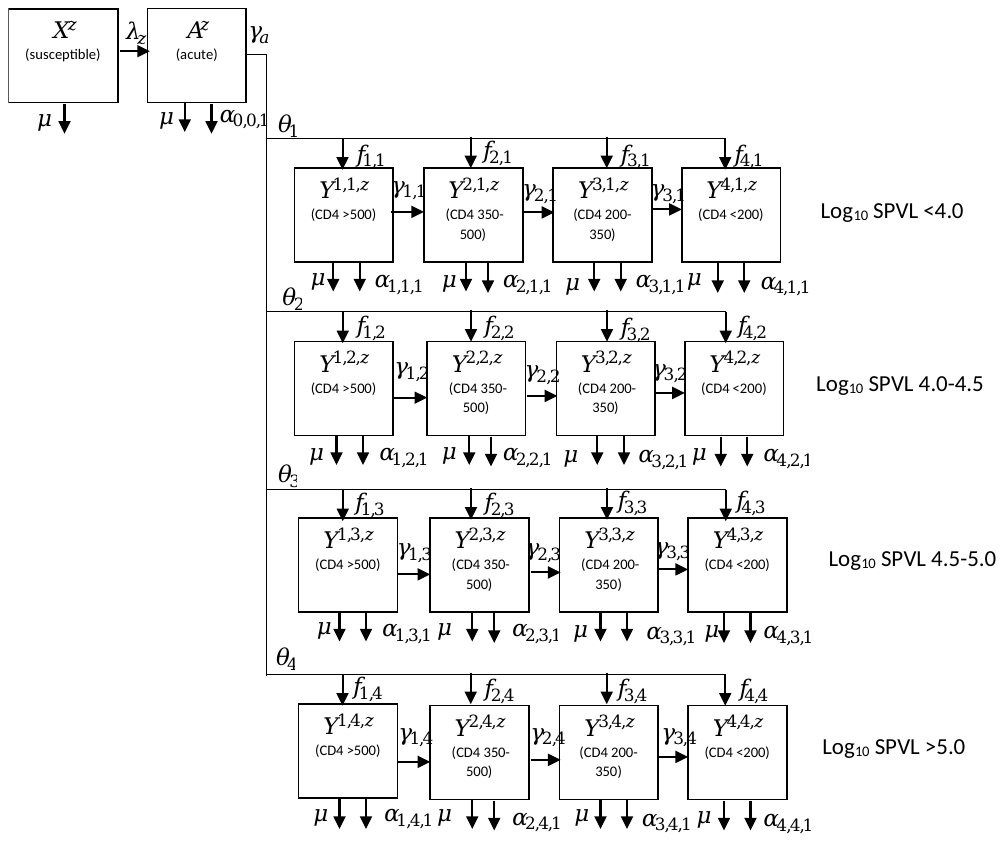

**Fig S2. HIV disease progression, by HIV states and SPVL, for those not on ART, and for those on ART but not adherent.** Superscripts on states and subscripts on HIV-related death rates are x,y,z (x = CD4 category; y = set-point viral load category; z = care state); subscripts for age and race are omitted for clarity.

Transitions between the different stages of care are shown in Fig S3. New men join the sexually active MSM population (through ageing into the population, sexual debut or immigration), at a rate $\Gamma$ and are assumed to all be uninfected with HIV initially. A proportion $p$ of new entrants are assumed to never routinely test for HIV and do not seek treatment until they become symptomatic (develop AIDS-defining illness); they enter the compartment for never-testing susceptibles ($X^{0}$). The remainder of new entrants enter compartment $X^{1}$, who are susceptibles who may undergo HIV testing. Susceptibles in either state may become infected at a rate $\lambda_{1}$. Susceptibles who may undergo testing can start PrEP at a per-capita rate $\delta\tau_{v,w,1}$, where $\tau_{v,w,1}$ is the age- and race-varying rate at which they test for HIV, and $\delta$ is the proportion of negative tests following which PrEP is offered and accepted. Those taking PrEP may drop out of PrEP at an age- and race-varying rate $\iota_{v,w}$, and return to the testing susceptible compartment ($X^{1}$).

Those never testing who become infected enter the infected compartments of never testers ($A^{0}/Y^{x,y,0},$ for acute/chronic infection, respectively). Those who may test enter the infected compartments of those undiagnosed but who may undergo HIV testing (${A^{1}/Y}^{x,y,1}$). Those on PrEP enter the infected on-PrEP compartments (${A^{2}/Y}^{x,y,2}$). Those on PrEP may drop out of PrEP at a rate $\iota$, moving to the equivalent disease stage for those who may test (${A^{1}/Y}^{x,y,1}$). Those who may test and those on PrEP undergo HIV testing at a per-capita rate $\tau_{v,w,z}$ – those on PrEP test at a higher rate, $\tau_{2}$, than those not on PrEP ($\tau_{1}$), and testing rates for those not on PrEP vary with age and race. A proportion $q$ of those testing are rapidly linked into care and move into the ‘in care’ compartments$A^{4}/Y^{x,y,4}$, the remainder (1-$q$) move into the ‘diagnosed not linked into care’ compartment (${A^{3}/Y}^{x,y,3}$). Those who are diagnosed but not in care can be linked into care, moving into compartments $A^{4}/Y^{x,y,4}$ at a rate $\epsilon_{v,w}$, and those linked into care may drop out from pre-ART care and go into the ‘diagnosed not linked into care’ compartment (${A^{3}/Y}^{x,y,3}$) at a rate ${\omega_{w}\phi_{5}}$ ($\phi_{5}$ is the rate of dropout from ART in the first year of treatment, $\omega_{w}$is the race-specific ratio of dropout from care relative to rate of dropout from ART). Those linked into care may begin ART, at a rate related to their CD4 count, $\xi_{x}$, with a proportion ($\chi$) who are adherent to their treatment moving into the first ART compartment,$A^{5}/$ $Y^{x,y,6}$, and those who are non-adherent (1-$\chi$) moving into compartment , $Y^{x,y,9}$. People at any other stage of the care continuum may also begin ART due to becoming symptomatic and seeking medical care, at a rate $\psi_{x,z}$, which is based upon CD4-count specific rates of incidence of AIDS-defining illness, and whether or not they have previously taken ART, and also move into the first ART compartment if they are adherent (proportion $\chi$), or the “on ART but not adherent” compartment ($Y^{x,y,9}$) if they are not adherent. Those in the non-adherent ART compartment are assumed to be fully infectious and have no survival benefit from ART, and progress in the same way as those not on ART.

People in the first ART compartment,$A^{5}/$ $Y^{x,y,5}$, are assumed to be partially virally suppressed, and they leave this compartment at a rate $\sigma_{y}$, where 1/$\sigma_{y}$ is the average duration from ART initiation to achieving viral suppression. $\sigma_{y}$ varies by SPVL, but not by initial CD4 count [1]. They move into the first fully virally suppressed compartment ($Y^{x,y,6}$), where they stay for the remainder of their first year on ART, and move into the next ART compartment (2^nd^ year;$Y^{x,y,7}$) at a rate $\eta_{y}$, where 1/$\eta_{y}$ (the average duration spent in the first year compartment) is estimated as 1- 1/$\sigma_{y}$. People move from the 2^nd^ year on ART compartment ($Y^{x,y,7}$) into the >2 years on ART compartment ($Y^{x,y,8}$) at a rate 1/year. The final fully suppressed compartment ($Y^{x,y,8}$) contains those who have remained on ART for more than 2 years and are still virally suppressed. For those on ART, the additional rate of HIV–related death from each of these compartments ($\alpha_{x,y,z}$) varies by CD4 count at ART initiation and duration on ART.

Those in any of the ART compartments may drop out of treatment at a rate $\phi_{z}$, which varies with time since initiation of ART. Dropouts from ART go initially into the dropout compartments, $Y^{x,y,10}$, where they progress through different CD4 compartments in the same way as those never on ART. Those dropping out of the adherent ART compartments (${A^{5},Y}^{x,y,5}-Y^{x,y,8})$, move into the same CD4 compartment as the one they were in when they started ART , those dropping out of the non-adherent ART compartment ($Y^{x,y,9}$) retain the CD4 count they had at the point of dropout. People remain in the same SPVL category after dropping out of ART. ART dropouts may re-initiate treatment due to developing AIDS symptoms and seeking medical care, at a rate $\psi_{x,10}$, or may re-enrol in HIV care, at a rate $\zeta$. Those re-entering care are not distinguished from those entering care for the first time. Likewise, those re-initiating treatment progress in the same way as those beginning ART for the first time, and are not distinguished from them.

We do not explicitly model individuals on ART gaining and losing viral suppression over time, due to a lack of data, but we do capture overall levels of viral suppression as well as dynamic (re-)entry and dropout from care and treatment.

The model was expressed as a set of differential equations which were solved numerically using a variable-stepsize eighth-order Runge-Kutta method ^2^.

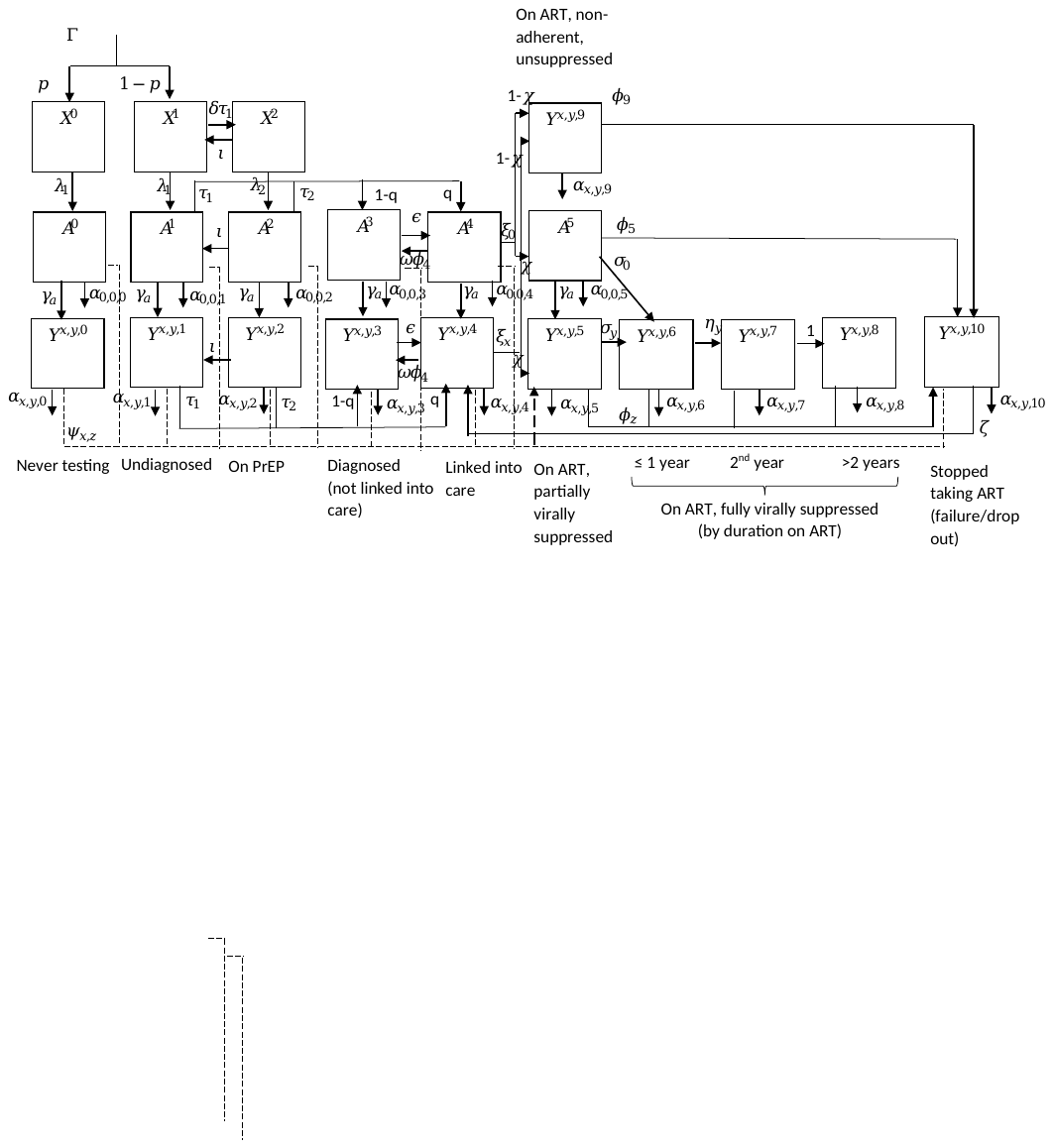

**Fig S3: Different stages of HIV care and transitions between them**

**Model equations**

*MSM who never get tested for HIV:*

$$\frac{d}{dt}\left( X_{v,w}^{0} \right)=\Gamma pm_{v,w}+{v\pi}_{w}X_{1-v,w}^{0}-X_{v,w}^{0}\left( \lambda_{v,w,1}+\mu_{v,w}+\left( 1-v \right)\pi_{w} \right)$$

$$\frac{d}{dt}\left( A_{v,w}^{0} \right)=\lambda_{v,w,1}X_{v,w}^{0}+{v\pi}_{w}A_{1-v,w}^{0}-A_{v,w}^{0}\left( \gamma_{a}+\mu_{v,w}+\alpha_{0,0,0}+\left( 1-v \right)\pi_{w}+\psi_{0,0} \right)$$

$$\frac{d}{dt}\left( Y_{v,w}^{1,y,0} \right)=\gamma_{a}{\theta_{y}f_{1,y}A}_{v,w}^{0}+{v\pi}_{w}Y_{1-v,w}^{1,y,0}-Y_{v,w}^{1,y,0}\left( \gamma_{1,y}+\mu_{v,w}+\alpha_{1,y,0}+\left( 1-v \right)\pi_{w}+\psi_{1,0} \right)$$

$$\frac{d}{dt}\left( Y_{v,w}^{x,y,0} \right)=\gamma_{a}{\theta_{y}f_{x,y}A}_{v,w}^{0}+\gamma_{x-1,y}Y_{v,w}^{x-1,y,0}+{v\pi}_{w}Y_{1-v,w}^{x,y,0}-Y_{v,w}^{x,y,0}\left( \gamma_{x,y}+\mu_{v,w}+\alpha_{x,y,0}+\left( 1-v \right)\pi_{w}+\psi_{x,0} \right);x\in\left\{ 2,3 \right\}$$

$$\frac{d}{dt}\left( Y_{v,w}^{4,y,0} \right)=\gamma_{a}{\theta_{y}f_{4,y}A}_{v,w}^{0}+\gamma_{3,y}Y_{v,w}^{3,y,0}+{v\pi}_{w}Y_{1-v,w}^{4,y,0}-Y_{v,w}^{4,y,0}\left( \mu_{v,w}+\alpha_{4,y,0}+\left( 1-v \right)\pi_{w}+\psi_{4,0} \right)$$

*MSM who may get tested, not diagnosed:*

$$\frac{d}{dt}\left( X_{v,w}^{1} \right)=\Gamma\left( 1-p \right)m_{v,w}+{v\pi}_{w}X_{1-v,w}^{1}+\iota_{v,w}X_{v,w}^{2}-X_{v,w}^{1}\left( \lambda_{v,w,1}+\mu_{v,w}+\left( 1-v \right)\pi_{w}+\delta\tau_{v,w,1} \right)$$

$$\frac{d}{dt}\left( A_{v,w}^{1} \right)=\lambda_{v,w,1}X_{v,w}^{1}+{v\pi}_{w}A_{1-v,w}^{1}+\iota_{v,w}A_{v,w}^{2}-A_{v,w}^{1}\left( \gamma_{a}+\mu_{v,w}+\alpha_{0,0,1}+\left( 1-v \right)\pi_{w}+\psi_{0,1}+\tau_{v,w,1} \right)$$

$$\frac{d}{dt}\left( Y_{v,w}^{1,y,1} \right)=\gamma_{a}{\theta_{y}f_{1,y}A}_{v,w}^{1}+{v\pi}_{w}Y_{1-v,w}^{1,y,1}+\iota_{v,w}Y_{v,w}^{1,y,2}-Y_{v,w}^{1,y,1}\left( \gamma_{1,y}+\mu_{v,w}+\alpha_{1,y,1}+\left( 1-v \right)\pi_{w}+\psi_{1,1}+\tau_{v,w,1} \right)$$

$$\frac{d}{dt}\left( Y_{v,w}^{x,y,1} \right)=\gamma_{a}{\theta_{y}f_{x,y}A}_{v,w}^{1}+\gamma_{x-1,y}Y_{v,w}^{x-1,y,1}+{v\pi}_{w}Y_{1-v,w}^{x,y,1}+\iota_{v,w}Y_{v,w}^{x,y,2}-Y_{v,w}^{x,y,1}\left( \gamma_{x,y}+\mu_{v,w}+\alpha_{x,y,1}+\left( 1-v \right)\pi_{w}+\psi_{x,1}+\tau_{v,w,1} \right);x\in\left\{ 2,3 \right\}$$

$$\frac{d}{dt}\left( Y_{v,w}^{4,y,1} \right)=\gamma_{a}{\theta_{y}f_{4,y}A}_{v,w}^{1}+\gamma_{3,y}Y_{v,w}^{3,y,1}+{v\pi}_{w}Y_{1-v,w}^{4,y,1}+\iota_{v,w}Y_{v,w}^{4,y,2}-Y_{v,w}^{4,y,1}\left( \mu_{v,w}+\alpha_{4,y,1}+\left( 1-v \right)\pi_{w}+\psi_{4,1}+\tau_{v,w,1} \right)$$

*MSM taking PrEP:*

$$\frac{d}{dt}\left( X_{v,w}^{2} \right)={\delta\tau_{v,w,1}X_{v,w}^{1}+v\pi}_{w}X_{1-v,w}^{2}-X_{v,w}^{2}\left( \lambda_{v,w,2}+\mu_{v,w}+\left( 1-v \right)\pi_{w}+\iota_{v,w} \right)$$

$$\frac{d}{dt}\left( A_{v,w}^{2} \right)=\lambda_{v,w,2}X_{v,w}^{2}+{v\pi}_{w}A_{1-v,w}^{2}-A_{v,w}^{2}\left( \gamma_{a}+\mu_{v,w}+\alpha_{0,0,2}+\left( 1-v \right)\pi_{w}+\psi_{0,2}+\iota_{v,w}+\tau_{v,w,2} \right)$$

$$\frac{d}{dt}\left( Y_{v,w}^{1,y,2} \right)=\gamma_{a}{\theta_{y}f_{1,y}A}_{v,w}^{2}+{v\pi}_{w}Y_{1-v,w}^{1,y,2}-Y_{v,w}^{1,y,2}\left( \gamma_{1,y}+\mu_{v,w}+\alpha_{1,y,2}+\left( 1-v \right)\pi_{w}+\psi_{1,2}+\iota_{v,w}+\tau_{v,w,2} \right)$$

$$\frac{d}{dt}\left( Y_{v,w}^{x,y,2} \right)=\gamma_{a}{\theta_{y}f_{x,y}A}_{v,w}^{2}+\gamma_{x-1,y}Y_{v,w}^{x-1,y,2}+{v\pi}_{w}Y_{1-v,w}^{x,y,2}-Y_{v,w}^{x,y,2}\left( \gamma_{x,y}+\mu_{v,w}+\alpha_{x,y,2}+\left( 1-v \right)\pi_{w}+\psi_{x,2}+\iota_{v,w}+\tau_{v,w,2} \right);x\in\left\{ 2,3 \right\}$$

$$\frac{d}{dt}\left( Y_{v,w}^{4,y,2} \right)=\gamma_{a}{\theta_{y}f_{4,y}A}_{v,w}^{2}+\gamma_{3,y}Y_{v,w}^{3,y,2}+{v\pi}_{w}Y_{1-v,w}^{4,y,2}-Y_{v,w}^{4,y,2}\left( \mu_{v,w}+\alpha_{4,y,2}+\left( 1-v \right)\pi_{w}+\psi_{4,2}+\iota_{v,w}+\tau_{v,w,2} \right)$$

*MSM diagnosed but not in care:*

$$\frac{d}{dt}\left( A_{v,w}^{3} \right)={v\pi}_{w}A_{1-v,w}^{3}+\left( 1-q \right)\left( \tau_{v,w,1}A_{v,w}^{1}+\tau_{v,w,2}A_{v,w}^{2} \right)+\omega_{w}\phi_{5}A_{v,w}^{4}-A_{v,w}^{3}\left( \gamma_{a}+\mu_{v,w}+\alpha_{0,0,3}+\left( 1-v \right)\pi_{w}+\psi_{0,3}+\epsilon_{w} \right)$$

$$\frac{d}{dt}\left( Y_{v,w}^{1,y,3} \right)=\gamma_{a}{\theta_{y}f_{1,y}A}_{v,w}^{3}+{v\pi}_{w}Y_{1-v,w}^{1,y,3}+\left( 1-q \right)\left( \tau_{v,w,1}Y_{v,w}^{1,y,1}+\tau_{v,w,2}Y_{v,w}^{1,y,2} \right)+\omega_{w}\phi_{5}Y_{v,w}^{1,y,4}{-Y}_{v,w}^{1,y,3}\left( \gamma_{1,y}+\mu_{v,w}+\alpha_{1,y,3}+\left( 1-v \right)\pi_{w}+\psi_{1,3}+\epsilon_{w} \right)$$

$$\frac{d}{dt}\left( Y_{v,w}^{x,y,3} \right)=\gamma_{a}{\theta_{y}f_{x,y}A}_{v,w}^{3}+\gamma_{x-1,y}Y_{v,w}^{x-1,y,3}+{v\pi}_{w}Y_{1-v,w}^{x,y,3}+\left( 1-q \right)\left( \tau_{v,w,1}Y_{v,w}^{x,y,1}+\tau_{v,w,2}Y_{v,w}^{x,y,2} \right)+\omega_{w}\phi_{5}Y_{v,w}^{x,y,4}-Y_{v,w}^{x,y,3}\left( \gamma_{x,y}+\mu_{v,w}+\alpha_{x,y,3}+\left( 1-v \right)\pi_{w}+\psi_{x,3}+\epsilon_{w} \right);x\in\left\{ 2,3 \right\}$$

$$\frac{d}{dt}\left( Y_{v,w}^{4,y,3} \right)=\gamma_{a}{\theta_{y}f_{4,y}A}_{v,w}^{3}+\gamma_{3,y}Y_{v,w}^{3,y,3}+{v\pi}_{w}Y_{1-v,w}^{4,y,3}+\left( 1-q \right)\left( \tau_{v,w,1}Y_{v,w}^{4,y,1}+\tau_{v,w,2}Y_{v,w}^{4,y,2} \right)+\omega_{w}\phi_{4}Y_{v,w}^{4,y,3}-Y_{v,w}^{4,y,2}\left( \mu_{v,w}+\alpha_{4,y,2}+\left( 1-v \right)\pi_{w}+\psi_{4,2}+\epsilon_{w} \right)$$

*MSM in care:*

$$\frac{d}{dt}\left( A_{v,w}^{4} \right)={v\pi}_{w}A_{1-v,w}^{4}+q\left( \tau_{v,w,1}A_{v,w}^{1}+\tau_{v,w,2}A_{v,w}^{2} \right)+\epsilon_{w}A_{v,w}^{3}-A_{v,w}^{4}\left( \gamma_{a}+\mu_{v,w}+\alpha_{0,0,4}+\left( 1-v \right)\pi_{w}+\psi_{0,4}+\omega_{w}\phi_{5}+\xi_{0} \right)$$

$$\frac{d}{dt}\left( Y_{v,w}^{1,y,4} \right)=\gamma_{a}{\theta_{y}f_{1,y}A}_{v,w}^{4}+{v\pi}_{w}Y_{1-v,w}^{1,y,4}+q\left( \tau_{v,w,1}Y_{v,w}^{1,y,1}+\tau_{v,w,2}Y_{v,w}^{1,y,2} \right)+\epsilon_{w}Y_{v,w}^{1,y,3}+\zeta Y_{v,w}^{1,y,10}-Y_{v,w}^{1,y,4}\left( \gamma_{1,y}+\mu_{v,w}+\alpha_{1,y,4}+\left( 1-v \right)\pi_{w}+\psi_{1,4}+\omega_{w}\phi_{5}+\xi_{1} \right)$$

$$\frac{d}{dt}\left( Y_{v,w}^{x,y,4} \right)=\gamma_{a}{\theta_{y}f_{x,y}A}_{v,w}^{4}+\gamma_{x-1,y}Y_{v,w}^{x-1,y,4}+{v\pi}_{w}Y_{1-v,w}^{x,y,4}+q\left( \tau_{v,w,1}Y_{v,w}^{x,y,1}+\tau_{v,w,2}Y_{v,w}^{x,y,2} \right)+\epsilon_{w}Y_{v,w}^{x,y,3}+\zeta Y_{v,w}^{x,y,10}-Y_{v,w}^{x,y,4}\left( \gamma_{x,y}+\mu_{v,w}+\alpha_{x,y,4}+\left( 1-v \right)\pi_{w}+\psi_{x,4}+\omega_{w}\phi_{5}+\xi_{x} \right);x\in\left\{ 2,3 \right\}$$

$$\frac{d}{dt}\left( Y_{v,w}^{4,y,4} \right)=\gamma_{a}{\theta_{y}f_{4,y}A}_{v,w}^{4}+\gamma_{3,y}Y_{v,w}^{3,y,4}+{v\pi}_{w}Y_{1-v,w}^{4,y,4}+q\left( \tau_{v,w,1}Y_{v,w}^{4,y,1}+\tau_{v,w,2}Y_{v,w}^{4,y,2} \right)+\epsilon_{w}Y_{v,w}^{4,y,3}+\zeta Y_{v,w}^{4,y,10}-Y_{v,w}^{4,y,4}\left( \mu_{v,w}+\alpha_{4,y,4}+\left( 1-v \right)\pi_{w}+\psi_{4,4}+\omega_{w}\phi_{5}+\xi_{4} \right)$$

*MSM on ART and adherent:*

$$\frac{d}{dt}\left( A_{v,w}^{5} \right)={v\pi}_{w}A_{1-v,w}^{5}+{\chi\xi}_{0}A_{v,w}^{4}+\sum_{Z=0}^{Z=4} {{\chi\psi}_{0,Z}A}_{v,w}^{Z}-A_{v,w}^{5}\left( \gamma_{a}+\mu_{v,w}+\alpha_{0,0,5}+\left( 1-v \right)\pi_{w}+\sigma_{0}+\phi_{5} \right)$$

$$\frac{d}{dt}\left( Y_{v,w}^{x,y,5} \right)={\gamma_{a}{\theta_{y}f_{x,y}A}_{v,w}^{5}+v\pi}_{w}Y_{1-v,w}^{x,y,5}+\chi\xi_{x}Y_{v,w}^{x,y,4}+\sum_{Z=0}^{Z=4} \chi{\psi_{x,Z}Y}_{v,w}^{x,y,4}+\chi\psi_{x,10}Y_{v,w}^{x,y,10}-Y_{v,w}^{x,y,5}\left( \mu_{v,w}+\alpha_{x,y,5}+\left( 1-v \right)\pi_{w}+\sigma_{y}+\phi_{5} \right)$$

$$\frac{d}{dt}\left( Y_{v,w}^{x,y,6} \right)={v\pi}_{w}Y_{1-v,w}^{x,y,6}+\sigma_{0}\theta_{y}f_{x,y}A_{v,w}^{5}{+\sigma_{y}Y}_{v,w}^{x,y,5}-Y_{v,w}^{x,y,6}\left( \mu_{v,w}+\alpha_{x,y,6}+\left( 1-v \right)\pi_{w}+\eta_{y}+\phi_{6} \right)$$

$$\frac{d}{dt}\left( Y_{v,w}^{x,y,7} \right)={v\pi}_{w}Y_{1-v,w}^{x,y,7}+\eta_{y}Y_{v,w}^{x,y,6}-Y_{v,w}^{x,y,7}\left( \mu_{v,w}+\alpha_{x,y,7}+\left( 1-v \right)\pi_{w}+1+\phi_{7} \right)$$

$$\frac{d}{dt}\left( Y_{v,w}^{x,y,8} \right)={v\pi}_{w}Y_{1-v,w}^{x,y,8}+Y_{v,w}^{x,y,7}-Y_{v,w}^{x,y,8}\left( \mu_{v,w}+\alpha_{x,y,8}+\left( 1-v \right)\pi_{w}+\phi_{8} \right)$$

*MSM on ART but non-adherent:*

$$\frac{d}{dt}\left( Y_{v,w}^{1,y,9} \right)=(1-{\chi)\xi}_{0}\theta_{y}f_{1,y}A_{v,w}^{4}+\left( 1-\chi\right)\xi_{1}Y_{v,w}^{1,y,4}+\sum_{Z=0}^{Z=4} {{\left( 1-\chi\right)\psi}_{0,Z}\theta_{y}f_{1,y}A}_{v,w}^{Z}+\sum_{Z=0}^{Z=4} \left( 1-\chi\right){\psi_{1,Z}Y}_{v,w}^{1,y,Z}+(1-\chi)\psi_{x,10}Y_{v,w}^{1,y,10}+{v\pi}_{w}Y_{1-v,w}^{1,y,9}-Y_{v,w}^{1,y,9}\left( \gamma_{1,y}+\mu_{v,w}+\alpha_{1,y,9}+\left( 1-v \right)\pi_{w}+\phi_{9} \right)$$

$$\frac{d}{dt}\left( Y_{v,w}^{x,y,9} \right)={(1-{\chi)\xi}_{0}\theta_{y}f_{x,y}A_{v,w}^{4}+\left( 1-\chi\right)\xi_{1}Y_{v,w}^{x,y,4}+\sum_{Z=0}^{Z=4} {{\left( 1-\chi\right)\psi}_{0,Z}\theta_{y}f_{x,y}A}_{v,w}^{Z}+\sum_{Z=0}^{Z=4} \left( 1-\chi\right){\psi_{x,Z}Y}_{v,w}^{x,y,Z}+(1-\chi)\psi_{x,10}Y_{v,w}^{x,y,10}+v\pi}_{w}Y_{1-v,w}^{x,y,9}+\gamma_{x-1,y}Y_{v,w}^{x-1,y,9}-Y_{v,w}^{x,y,9}\left( \gamma_{x,y}+\mu_{v,w}+\alpha_{x,y,9}+\left( 1-v \right)\pi_{w}+\phi_{9} \right);x\in\left\{ 2,3 \right\}$$

$$\frac{d}{dt}\left( Y_{v,w}^{4,y,9} \right)=(1-{\chi)\xi}_{0}\theta_{y}f_{4,y}A_{v,w}^{4}+\left( 1-\chi\right)\xi_{1}Y_{v,w}^{4,y,4}+\sum_{Z=0}^{Z=4} {{\left( 1-\chi\right)\psi}_{0,Z}\theta_{y}f_{4,y}A}_{v,w}^{Z}+\sum_{Z=0}^{Z=4} \left( 1-\chi\right){\psi_{x,Z}Y}_{v,w}^{4,y,Z}+(1-\chi)\psi_{x,10}Y_{v,w}^{4,y,10}+{v\pi}_{w}Y_{1-v,w}^{4,y,9}+\gamma_{3}Y_{v,w}^{3,y,9}-Y_{v,w}^{4,y,9}\left( \mu_{v,w}+\alpha_{4,y,9}+\left( 1-v \right)\pi_{w}+\phi_{9} \right)$$

*MSM dropped out of ART:*

$$\frac{d}{dt}\left( Y_{v,w}^{1,y,10} \right)={\sum_{Z=5}^{Z=9} {\phi_{Z}Y}_{v,w}^{1,y,Z}+v\pi}_{w}Y_{1-v,w}^{1,y,10}+\theta_{y}f_{x,y}\phi_{5}A_{v,w}^{5}-Y_{v,w}^{1,y,10}\left( \gamma_{1,y}+\mu_{v,w}+\alpha_{1,y,10}+\left( 1-v \right)\pi_{w}+\psi_{1,10}+\zeta\right)$$

$$\frac{d}{dt}\left( Y_{v,w}^{x,y,10} \right)=\sum_{Z=5}^{Z=9} {\phi_{Z}Y}_{v,w}^{x,y,Z}{+v\pi}_{w}Y_{1-v,w}^{x,y,10}{+\theta}_{y}f_{x,y}\phi_{5}A_{v,w}^{5}+\gamma_{x-1,y}Y_{v,w}^{x-1,y,10}-Y_{v,w}^{x,y,10}\left( \gamma_{x,y}+\mu_{v,w}+\alpha_{x,y,10}+\left( 1-v \right)\pi_{w}+\psi_{x,10}+\zeta\right);x\in\left\{ 2,3 \right\}$$

$$\frac{d}{dt}\left( Y_{v,w}^{4,y,10} \right)=\sum_{Z=5}^{Z=9} {\phi_{Z}Y}_{v,w}^{4,y,Z}+{{v\pi}_{w}Y_{1-v,w}^{4,y,10}+\theta}_{y}f_{x,y}\phi_{5}A_{v,w}^{5}+\gamma_{3}Y_{v,w}^{3,y,10}-Y_{v,w}^{4,y,10}\left( \mu_{v,w}+\alpha_{4,y,10}+\left( 1-v \right)\pi_{w}+\psi_{4,10}+\zeta\right)$$

*Force of infection*

Not on PrEP:

$$\lambda_{v,w,1}=1-\left( {\prod_{j=1}^{j=3} \prod_{v^{'}=0}^{v^{'}=1} \prod_{w^{'}=0}^{w^{'}=1} \left( \frac{\sum_{z=0}^{z=2} \left( X_{v',w'}^{z} \right)}{N_{v',w'}}+\frac{\sum_{z=0}^{z=4} \left( A_{v',w'}^{z} \right)}{N_{v',w'}}\left( 1-d_{1}\beta\left( 1-e_{c}s_{c,j} \right)\left( 1-e_{n}s_{n} \right) \right)^{n_{j}}+\sum_{y=1}^{y=4} \left( {\frac{\sum_{x=1}^{x=3} \left( \sum_{z=0}^{z=4} \left( Y_{v',w'}^{x,y,z} \right)+Y_{v',w'}^{x,y,9}+Y_{v',w'}^{x,y,10} \right)}{N_{v',w'}}\left( 1-h_{y}\beta\left( 1-e_{c}s_{c,j} \right)\left( 1-e_{n}s_{n} \right) \right)}^{n_{j}} \right)+\sum_{y=1}^{y=4} \left( {\frac{\left( \sum_{z=0}^{z=4} \left( Y_{v',w'}^{4,y,z} \right)+Y_{v',w'}^{4,y,9}+Y_{v',w'}^{4,y,10} \right)}{N_{v',w'}}\left( 1-d_{2}h_{y}\beta\left( 1-e_{c}s_{c,j} \right)\left( 1-e_{n}s_{n} \right) \right)}^{n_{j}} \right)+\frac{A_{v',w'}^{5}}{N_{v',w'}}\left( 1-d_{3}\beta\left( 1-e_{c}s_{c,j} \right)\left( 1-e_{n}s_{n} \right) \right)^{n_{j}}+\sum_{y=1}^{y=4} \left( {\frac{\sum_{x=1}^{x=3} \left( Y_{v',w'}^{x,y,5} \right)}{N_{v',w'}}\left( 1-d_{4}h_{y}\beta\left( 1-e_{c}s_{c,j} \right)\left( 1-e_{n}s_{n} \right) \right)}^{n_{j}} \right)+\sum_{y=1}^{y=4} \left( {\frac{\left( Y_{v',w'}^{4,y,5} \right)}{N_{v',w'}}\left( 1-d_{5}h_{y}\beta\left( 1-e_{c}s_{c,j} \right)\left( 1-e_{n}s_{n} \right) \right)}^{n_{j}} \right)+\sum_{y=1}^{y=4} \left( {\frac{\sum_{x=1}^{x=4} \sum_{z=6}^{z=8} \left( Y_{v',w'}^{x,y,z} \right)}{N_{v',w'}}\left( 1-d_{6}h_{y}\beta\left( 1-e_{c}s_{c,j} \right)\left( 1-e_{n}s_{n} \right) \right)}^{n_{j}} \right) \right)}^{\rho_{vw,v'w',j}c_{v,w,j}} \right)$$

On PrEP:

$$\lambda_{v,w,1}=1-\left( {\prod_{j=1}^{j=3} \prod_{v^{'}=0}^{v^{'}=1} \prod_{w^{'}=0}^{w^{'}=1} \left( \frac{\sum_{z=0}^{z=2} \left( X_{v',w'}^{z} \right)}{N_{v',w'}}+\frac{\sum_{z=0}^{z=4} \left( A_{v',w'}^{z} \right)}{N_{v',w'}}\left( 1-d_{1}\beta\left( 1-e_{c}s_{c,j} \right)\left( 1-e_{n}s_{n} \right)(1-e_{p}s_{p,v,w}) \right)^{n_{j}}+\sum_{y=1}^{y=4} \left( {\frac{\sum_{x=1}^{x=3} \left( \sum_{z=0}^{z=4} \left( Y_{v',w'}^{x,y,z} \right)+Y_{v',w'}^{x,y,9}+Y_{v',w'}^{x,y,10} \right)}{N_{v',w'}}\left( 1-h_{y}\beta\left( 1-e_{c}s_{c,j} \right)\left( 1-e_{n}s_{n} \right)(1-e_{p}s_{p,v,w}) \right)}^{n_{j}} \right)+\sum_{y=1}^{y=4} \left( {\frac{\left( \sum_{z=0}^{z=4} \left( Y_{v',w'}^{4,y,z} \right)+Y_{v',w'}^{4,y,9}+Y_{v',w'}^{4,y,10} \right)}{N_{v',w'}}\left( 1-d_{2}h_{y}\beta\left( 1-e_{c}s_{c,j} \right)\left( 1-e_{n}s_{n} \right)(1-e_{p}s_{p,v,w}) \right)}^{n_{j}} \right)+\frac{A_{v',w'}^{5}}{N_{v',w'}}\left( 1-d_{3}\beta\left( 1-e_{c}s_{c,j} \right)\left( 1-e_{n}s_{n} \right)(1-e_{p}s_{p,v,w}) \right)^{n_{j}}+\sum_{y=1}^{y=4} \left( {\frac{\sum_{x=1}^{x=3} \left( Y_{v',w'}^{x,y,5} \right)}{N_{v',w'}}\left( 1-d_{4}h_{y}\beta\left( 1-e_{c}s_{c,j} \right)\left( 1-e_{n}s_{n} \right)(1-e_{p}s_{p,v,w}) \right)}^{n_{j}} \right)+\sum_{y=1}^{y=4} \left( {\frac{\left( Y_{v',w'}^{4,y,5} \right)}{N_{v',w'}}\left( 1-d_{5}h_{y}\beta\left( 1-e_{c}s_{c,j} \right)\left( 1-e_{n}s_{n} \right)(1-e_{p}s_{p,v,w}) \right)}^{n_{j}} \right)+\sum_{y=1}^{y=4} \left( {\frac{\sum_{x=1}^{x=4} \sum_{z=6}^{z=8} \left( Y_{v',w'}^{x,y,z} \right)}{N_{v',w'}}\left( 1-d_{6}h_{y}\beta\left( 1-e_{c}s_{c,j} \right)\left( 1-e_{n}s_{n} \right)(1-e_{p}s_{p,v,w}) \right)}^{n_{j}} \right) \right)}^{\rho_{vw,v'w',j}c_{v,w,j}} \right)$$

where the total number of MSM partners in age group $v'$ and race group $w'$ is calculated as:

$$N_{v',w'}=\sum_{z=0}^{z=1} \left( X_{v',w'}^{z} \right)+\sum_{z=0}^{z=4} \left( A_{v',w'}^{z} \right)+\sum_{x=1}^{x=4} \sum_{y=1}^{y=4} \sum_{z=0}^{z=9} \left( Y_{v',w'}^{x,y,z} \right)$$

Infection risk is estimated for three partner types (j = 1: regular partners, j = 2: casual partners; j = 3: commercial partners). $e_{c}$ is per-sex-act condom efficacy, $s_{c,j}$ is the proportion of sex acts in which a condom is used with partners of type $j$, $e_{n}$ is per-sex act reduction in HIV acquisition risk due to male circumcision, $s_{n}$ is the proportion of MSM who are circumcised, $e_{p}$ is the per-sex act reduction in HIV acquisition due to PrEP use, and $s_{p,v,w}$ is the proportion of men in each age and race group who are adherent to PrEP. $\beta$ is the average probability of acquiring HIV infection from an anal sex act with an HIV-positive male partner with chronic infection and CD4>200 cells/µl who is not taking ART,$\rho_{vw,v'w',j}$ is, for MSM in age group $v$ and race group $w$, the proportion of partners of type $j$ who are in age group $v'$ and race group $w'$ . $c_{v,w,j}$ is the average number of new partners per year of type $j$ for MSM in age group $v$ and race group $w$, $n_{j}$ is the average number of sex acts per partnership for a partnership of type $j$, $d_{1}$is the relative infectiousness of those in the acute versus chronic stage of infection, $d_{2}$ is the relative infectiousness of those with CD4<200 cells/µl versus those with chronic infection and CD4>200 cells/µl, $d_{3}, d_{4},d_{5}$are the relative infectiousness of those on ART with a partially suppressed viral load who have acute infection, chronic infection (CD4>200 cells/µl) or CD4<200 cells/µl, respectively, versus those untreated with chronic infection and CD4>200 cells/µl , $d_{6}$is the relative infectiousness of those on ART with a fully suppressed viral load versus those untreated with chronic infection and CD4>200 cells/µl, and $h_{y}$ is the relative infectiousness of those not fully virally suppressed who have SPVL $y$.

The relative infectiousness of those on ART with a partially suppressed viral load are calculated as follows:

$${d_{3}=d}_{6}+d_{r}\left( d_{1}-d_{6} \right)$$

$${d_{4}=d}_{6}+d_{r}\left( 1-d_{6} \right)$$

$${d_{5}=d}_{6}+d_{r}\left( d_{2}-d_{6} \right)$$

Where $d_{r}$ is the relative level of infectiousness of those partially suppressed, scaled between the level for those fully suppressed ${(d}_{r}=0)$ and those unsuppressed ($d_{r}=1)$.

**Calibrating the model to PrEP coverage**

Age- and race-specific rates of PrEP adherence and retention were assumed to be the same as for clinic-referred MSM in the US PrEP Demo project ^3,4^. We assumed PrEP use started in 2012, with PrEP initiation (the proportion of HIV-uninfected MSM initiating PrEP following routine HIV testing) increasing linearly up to 2020. The model was calibrated to 2014 and 2017 NHBS survey data on PrEP coverage ^5^ ^6^, by varying the final level of PrEP initiation (i.e. the proportion initiating PrEP following routine HIV testing in 2020), until PrEP coverage in the model lay between 40% and 100% of the survey data estimate (which was of any PrEP use in the preceding 12 months).

**Table S1. Parameters used in the HIV transmission model, with source and justification**

| **Symbol** | **Parameter** | **Range of values** | **Source/justification** |
| --- | --- | --- | --- |
| INITIAL CONDITIONS | | | |
| $N_{0}$ | Initial size of MSM population (1984) | 6765-8326 | 260,199 men aged 18+ in the 1980 Baltimore census; Purcell et al. 2012^7^ estimate % of US men had same-sex behaviour last 12 months 2.9% (95% CI 2.6-3.2%) |
|  | Percentage of MSM who are black in 1984 | 50-64 | Main estimate: overall population 1980 census. Upper limit: MSM in NHBS 2004; lower limit: lower 95% CI in NHBS 2004^1^ |
|  | Percentage of black MSM aged 18-24 in 1984 | 16-31 | Lower bound: black men in 2010 census  Upper bound: black MSM NHBS 2004 (upper 95% CI)^1^ |
|  | Percentage of white MSM aged 18-24 in 1984 | 14-28 | Lower bound: white men in 2010 census  Upper bound: white MSM NHBS 2004 (upper 95% CI)^1^ |
|  | HIV prevalence black MSM 1984 (%) | 15-44 | MACS baseline black MSM ^8^– lower bound a third of this as non-random sample |
|  | HIV prevalence white MSM 1984 (%) | 9-28 | MACS baseline white MSM ^8^– lower bound a third of this as non-random sample |
| *Demography* | |  |  |
| $\Gamma$ | Rate at which new MSM join the sexually active MSM population (per year) | 100-400 (fitting to census demography)  200-800 (fitting to NHBS demography) | estimate |
| $m_{black}$ | Percentage of new incoming MSM who are black | 60-85 | Baltimore census 1990-2010; NHBS 2004-2011 ^1^ |
| $m_{young,0}$ | Percentage of new incoming black MSM who are aged 18-24 years old | 72-87 | % of black MSM in NHBS who say they entered sexually active Baltimore MSM population aged <25 – 2008 & 2011 NHBS ^1^ |
| $m_{young,1}$ | Percentage of new incoming white MSM who are aged 18-24 years old | 50-71 (fitting to census demography)  37-71 (fitting to NHBS demography) | % of white MSM in NHBS who say they entered sexually active Baltimore MSM population aged <25 – 2008 & 2011 NHBS^1^ |
| $\pi_{0}$ | rate of moving from 18-24 year old age group to >24 year old age group, black MSM, per year | 0.17 (fixed) | Mean age at joining the local MSM population in NHBS 2008 and 2011 for 18-24 yr old MSM ~16 yrs old (95% CI 15-17) ^1^ |
| $\pi_{1}$ | rate of moving from 18-24 year old age group to >24 year old age group, white MSM, per year | 0.17-0.25 | Mean age at joining the local MSM population in NHBS 2008 and 2011 for 18-24 yr old MSM ~18/19 yrs old (95% CI 16/17-20) ^1^ |
| $\mu_{0,0}$ | Non-HIV related death rate, 18-24 year old black men, per year | 0.0011-0.0015 | CDC WONDER database data for Maryland; data for 15-24 years olds |
| $\mu_{1,0}$ | Non-HIV related death/leaving rate, >24 year old black men, per year | 0.011-0.04 (census fitting)  0.041-0.11 (NHBS fitting) | CDC WONDER database data for Maryland; average death rate over ages 26-64 years old  Upper bound: add on 1/36 (double current duration as an MSM)  NHBS fitting: additionally assume extra rate of ceasing to attend NHBS venues |
| $\mu_{0,1}$ | Non-HIV related death rate, 18-24 year old white men, per year | 0.00075-0.001 | CDC WONDER database data for Maryland; data for 15-24 years olds |
| $\mu_{1,1}$ | Non-HIV related death/leaving rate, >24 year old white men, per year | 0.033-0.1 (census fitting)  0.058-0.128 (NHBS fitting) | High rates reflecting out-migration plus rates of ceasing sexual activity;  NHBS fitting: additionally assume extra rate of ceasing to attend NHBS venues |
| *Sexual behaviour* | | | |
| $n_{1}$ | Number of sex acts per main partnership | 40-470 | 48.2-85.1 sex episodes/year with main partners ^9^, partnerships last 3.5-5.5 years ^10,11^, but assume some are shorter (~1 year) |
| $n_{2}$ | Number of sex acts per casual partnership | 1.5-6 | 3-4.9 sex episodes/year ^9^, partnerships last 0.5-1.3 years ^10^ |
| $n_{3}$ | Number of sex acts per commercial partnership | 1-2 | assumed |
| $c_{0,0,1}$ | Number of new main partners per year, 18-24 year old black MSM | 0.58-0.8 | NHBS 2004, 2008, 2011^1^ |
| $c_{0,0,2}$ | Number of new casual partners per year, 18-24 year old black MSM 2011 onwards^a^ | 1.54-2.09 | NHBS 2011^1^ |
| $c_{0,0,3}$ | Number of new commercial partners per year, 18-24 year old black MSM 2011 onwards^a^ | 0-1.36 | NHBS 2011^1^ |
| $c_{1,0,1}$ | Number of new main partners per year, >24 year old black MSM | 0.36-0.57 | NHBS 2004, 2008, 2011^1^ |
| $c_{1,0,2}$ | Number of new casual partners per year, >24 year old black MSM 2011 onwards^a^ | 0.81-1.24 | NHBS 2011^1^ |
| $c_{1,0,3}$ | Number of new commercial partners per year, >24 year old black MSM 2011 onwards^a^ | 0.15-0.85 | NHBS 2011 ^1^ |
| $c_{0,1,1}$ | Number of new main partners per year, 18-24 year old white MSM | 0.08-0.37 | NHBS 2004, 2008, 2011 ^1^ |
| $c_{0,1,2}$ | Number of new casual partners per year, 18-24 year old white MSM 2011 onwards^a^ | 0.05-0.93 | NHBS 2011^1^ |
| $c_{0,1,3}$ | Number of new commercial partners per year, 18-24 year old white MSM 2011 onwards^a^ | 0-0.28 | NHBS 2011^1^ |
| $c_{1,1,1}$ | Number of new main partners per year, >24 year old white MSM | 0.11-0.21 | NHBS 2004, 2008, 2011 ^1^ |
| $c_{1,1,2}$ | Number of new casual partners per year, >24 year old white MSM 2011 onwards^a^ | 0.28-1.07 | NHBS 2011^1^ |
| $c_{1,1,3}$ | Number of new commercial partners per year, >24 year old white MSM 2011 onwards^a^ | 0-0.07 | NHBS 2011^1^ |
| Partner_number_decline | absolute decline per year in the number of new casual or commercial partners | 0.17-0.36 | From trends in NHBS data on number of commercial and causal partners 2004-2011^1^ |
| Mixing parameter for age mixing | Scale between fully proportionate and fully assortative mixing by age | 0.25-0.35 | estimated from NHBS 2011 data on last partner^1^ |
| Mixing parameter for race mixing | Scale between fully proportionate and fully assortative mixing by race | 0.7-0.8 | 0.75 estimated from NHBS 2011 data on last partner and 0.74 from NHBS additional data 2008 ^1^ ^12^ |
| Early_condom_use | Minimum level of condom use at start of the HIV epidemic (% of sex acts) | 0-30 | No data |
| $s_{c,1,0}$ | Percentage of sex acts in which a condom is used, main partnerships where both partners are black, 2004 onwards^a^ | 47-67 | condom use last sex act reported by black MSM with main partners NHBS 2004-2011 ^1^ |
| $s_{c,1,1}$ | Percentage of sex acts in which a condom is used, main partnerships where one or both partners are white, 2004 onwards^a^ | 30-39 | condom use last sex act reported by white MSM with main partners NHBS 2004-2011^1^ |
| $s_{c,2}$ | Percentage of sex acts in which a condom is used, casual partnerships (any race partner), 2004 onwards^a^ | 63-72 | condom use last sex act reported in casual partnerships NHBS 2004-2011^1^ |
| $s_{c,3}$ | Percentage of sex acts in which a condom is used, commercial partnerships (any race partner), 2004 onwards^a^ | 21-78 | condom use last sex act reported in commercial partnerships NHBS 2004 & 2008^1^ |
| Condom_increase_1 | Yearly increase in % of sex acts in which condoms are used, all partnerships prior to 2008 | 2.4-4 | From trend in data from NHBS 2004-2008, averaging over condom use in main and casual partnerships^1^ |
| Condom_increase_2 | Yearly increase in % of sex acts in which condoms are used, all partnerships between 2008 and 2011 | -2.4-+0.2 | From trends in data from NHBS 2008-2011 and 2008-2014, averaging over condom use in main and casual partnerships^1^ |
| *HIV disease progression* | |  |  |
| $1/\gamma_{a}$ | Average duration of acute infection, months | 2-6 | ^13,14^ |
| $\alpha_{0,0,z}$ | HIV-related death rate for those with acute HIV infection, per year | 0 (fixed) | assumption |
| $\alpha_{1,y,0}$  $\alpha_{1,y,1}$  $\alpha_{1,y,2}$  $\alpha_{1,y,3}$  $\alpha_{1,y,8}$  $\alpha_{1,y,9}$ | HIV-related death rate for those with CD4>500, off ART, per year | 0.0009-0.0054 | aged 25-44 in the European CASCADE cohort ^15^; general population death rate subtracted |
| $\alpha_{2,y,0}$  $\alpha_{2,y,1}$  $\alpha_{2,y,2}$  $\alpha_{2,y,3}$ | HIV-related death rate for those with CD4 350-500, off ART, per year | 0.0009-0.0069 | aged 25-44 in the European CASCADE cohort ^15^; general population death rate subtracted |
| $\alpha_{3,y,1}$  $\alpha_{3,y,1}$ | HIV related death rate for those with CD4 200-350, off ART, per year | 0.0045-0.0135 | aged 25-44 in the European CASCADE cohort ^15^; general population death rate subtracted |
| 1/$\alpha_{4,1,1}$ | Inverse of HIV-related death rate for those with CD4<200, SPVL<4.0, off ART (years) | 3.28-12.87 | Netherlands ATHENA cohort ^16^ |
| 1/$\alpha_{4,2,1}$ | Inverse of HIV-related death rate for those with CD4<200, SPVL 4.0-4.5, off ART (years) | 1.43-6.09 | Netherlands ATHENA cohort ^16^ |
| 1/$\alpha_{4,3,1}$ | Inverse of HIV-related death rate for those with CD4<200, SPVL 4,5-5,0, off ART (years) | 4.41-23.64 | Netherlands ATHENA cohort ^16^ |
| 1/$\alpha_{4,4,1}$ | Inverse of HIV-related death rate for those with CD4<200, SPVL>5.0, off ART (years) | 1.32-3.59 | Netherlands ATHENA cohort ^16^ |
| $\alpha_{1,y,4}$,$\alpha_{2,y,4}$  $\alpha_{1,y,5}$,$\alpha_{2,y,5}$  $\alpha_{1,y,6}$,$\alpha_{2,y,6}$  $\alpha_{1,y,7}$  $\alpha_{2,y,7}$ | HIV-related mortality for those with CD4>500 or CD4 350-500 at start of treatment, for 1^st^ , 2^nd^ and subsequent years on ART, per year | 0-0.003 | From probabilities for those with CD4>350 ^17^; general population death rate subtracted ^18^ |
| $a_{1}$ | Relative mortality of those with CD4 200-350 vs CD4>350 at start of treatment, 1^st^ year on ART | 1.2-2.8 | ^19^ |
| $a_{2}$ | Relative mortality of those with CD4 200-350 vs CD4>350 at start of treatment, 2^nd^ year on ART | 1-2.2 | ^19^ Upper limit reduced to give main estimate as midpoint |
| $a_{3}$ | Relative mortality of those with CD4 200-350 vs CD4>350 at start of treatment, 3^rd^ year + on ART | 1-1.4 | ^19^ Upper limit reduced to give main estimate as midpoint |
| $a_{4}$ | Relative mortality of those with CD4 <200 vs CD4>350 at start of treatment, 1^st^ year on ART | 1.8-5.2 | ^19^ Main estimate and lower bound: CD4 100-199; upper bound from those with CD4 25-49 |
| $a_{5}$ | Relative mortality of those with CD4 <200 vs CD4>350 at start of treatment, 2^nd^ year on ART | 1.3-6.2 | ^19^ Main estimate and lower bound: CD4 100-199; upper bound from those with CD4 25-49 |
| $a_{6}$ | Relative mortality of those with CD4 <200 vs CD4>350 at start of treatment, 3^rd^ year + on ART | 1-3.2 | ^19^ Main estimate and lower bound: CD4 100-199; upper bound from those with CD4 50-99 |
| $b_{1}$ | Relative mortality of those with AIDS before ART initiation vs without, 1^st^ year on ART | 3.0-4.8 | ^19^ |
| $b_{2},b_{3}$ | Relative mortality of those with AIDS before ART initiation vs without, 2^nd^ , 3^rd^ + years on ART | 1.4-2.6 | ^19^ |
| $k_{4}$ | Percentage of those starting ART with CD4<200 who have a prior AIDS diagnosis | 40-60 | ^20^ |
| $\theta_{2}$ | Percentage of HIV-positive MSM with a SPVL 4.0-4.5 | 25 (fixed) | Netherlands ATHENA cohort ^16^; US MSM (MACS cohort)^21,22^ |
| $\theta_{3}$ | Percentage of HIV-positive MSM with a SPVL 4.5-5.0 | 25-40 | Netherlands ATHENA cohort ^16^; US MSM (MACS cohort)^21,22^ |
| $\theta_{4}$ | Percentage of HIV-positive MSM with a SPVL >5.0 | 10-25 | Netherlands ATHENA cohort ^16^; US MSM (MACS cohort)^22^ |
| ${1/\gamma}_{1,1}$ | Average duration spent with CD4>500 cells/µl, for those with SPVL <4.0 (years) | 4.56-6.37 | Netherlands ATHENA cohort ^16^ |
| ${1/\gamma}_{2,1}$ | Average duration spent with CD4 350-500, for those with SPVL <4.0 (years) | 2.98-4.53 | Netherlands ATHENA cohort ^16^ |
| ${1/\gamma}_{3,1}$ | Average duration spent with CD4 200-350, for those with SPVL <4.0 (years) | 5.04-13.69 | Netherlands ATHENA cohort ^16^ |
| ${1/\gamma}_{1,2}$ | Average duration spent with CD4>500, for those with SPVL 4.0-4.5 (years) | 2.68-3.64 | Netherlands ATHENA cohort ^16^ |
| ${1/\gamma}_{2,2}$ | Average duration spent with CD4 350-500, for those with SPVL 4.0-4.5 (years) | 2.65-3.64 | Netherlands ATHENA cohort ^16^ |
| ${1/\gamma}_{3,2}$ | Average duration spent with CD4 200-350, for those with SPVL 4.0-4.5 (years) | 5.46-15.55 | Netherlands ATHENA cohort ^16^ |
| ${1/\gamma}_{1,3}$ | Average duration spent with CD4>500, for those with SPVL 4.5-5.0 (years) | 2.08-2.64 | Netherlands ATHENA cohort ^16^ |
| ${1/\gamma}_{2,3}$ | Average duration spent with CD4 350-500, for those with SPVL 4.5-5.0 (years) | 1.98-2.72 | Netherlands ATHENA cohort ^16^ |
| ${1/\gamma}_{3,3}$ | Average duration spent with CD4 200-350, for those with SPVL 4.5-5.0 (years) | 4.73-10.22 | Netherlands ATHENA cohort ^16^ |
| ${1/\gamma}_{1,4}$ | Average duration spent with CD4>500, for those with SPVL ≥5.0 (years) | 1.28-1.76 | Netherlands ATHENA cohort ^16^ |
| ${1/\gamma}_{2,4}$ | Average duration spent with CD4 350-500, for those with SPVL ≥5.0 (years) | 1.22-1.69 | Netherlands ATHENA cohort ^16^ |
| ${1/\gamma}_{3,4}$ | Average duration spent with CD4 200-350, for those with SPVL ≥5.0 (years) | 2.12-4.19 | Netherlands ATHENA cohort ^16^ |
| ${1/\sigma}_{0}$ | Average duration from ART initiation to viral suppression (VL < 200 copies/ml) for those with acute HIV infection (months) | 3.93-8.50 | Pregnant women, Kenya^23^ |
| ${1/\sigma}_{1}$ | Average duration from ART initiation to viral suppression (VL < 200 copies/ml) for those with log_10_ SPVL <4.0 (months) | 0.95-4.1 | Data from Johns Hopkins (Baltimore) and Fenway (Boston); estimate is weighted average of median values from 2 sites^1^ |
| ${1/\sigma}_{2}$ | Average duration from ART initiation to viral suppression (VL < 200 copies/ml) for those with log_10_ SPVL 4.0-4.5 (months) | 1.03-4.75 | Data from Johns Hopkins (Baltimore) and Fenway (Boston); estimate is weighted average of median values from 2 sites^1^ |
| ${1/\sigma}_{3}$ | Average duration from ART initiation to viral suppression (VL < 200 copies/ml) for those with log_10_ SPVL 4.5-5.0 (months) | 1.4-6.43 | Data from Johns Hopkins (Baltimore) and Fenway (Boston); estimate is weighted average of median values from 2 sites^1^ |
| ${1/\sigma}_{4}$ | Average duration from ART initiation to viral suppression (VL < 200 copies/ml) for those with log_10_ SPVL >5.0 (months) | 2.03-6.49 | Data from Johns Hopkins (Baltimore) and Fenway (Boston); estimate is weighted average of median values from 2 sites^1^ |
| $f_{1,1}$ | Percentage with CD4 >500 after seroconversion, for those with SPVL <4.0 | 81-91 | Netherlands ATHENA cohort ^16^ |
| $f_{3,1}$ | Percentage with CD4 200-350 after seroconversion, for those with SPVL <4.0 | 0-4 | Netherlands ATHENA cohort ^16^ |
| $f_{4,1}$ | Percentage with CD4 <200 after seroconversion, for those with SPVL <4.0 | 0 (fixed) | Netherlands ATHENA cohort ^16^ |
| $f_{1,2}$ | Percentage with CD4 >500 after seroconversion, for those with SPVL 4.0-4.5 | 72-83 | Netherlands ATHENA cohort ^16^ |
| $f_{3,2}$ | Percentage with CD4 200-350 after seroconversion, for those with SPVL 4.0-4.5 | 1-5 | Netherlands ATHENA cohort ^16^ |
| $f_{4,2}$ | Percentage with CD4 <200 after seroconversion, for those with SPVL 4.0-4.5 | 0 (fixed) | Netherlands ATHENA cohort ^16^ |
| $f_{1,3}$ | Percentage with CD4 >500 after seroconversion, for those with SPVL 4.5-5.0 | 69-79 | Netherlands ATHENA cohort ^16^ |
| $f_{3,3}$ | Percentage with CD4 200-350 after seroconversion, for those with SPVL 4.5-5.0 | 3-8 | Netherlands ATHENA cohort ^16^ |
| $f_{4,3}$ | Percentage with CD4 <200 after seroconversion, for those with SPVL 4.5-5.0 | 0 (fixed) | Netherlands ATHENA cohort ^16^ |
| $f_{1,4}$ | Percentage with CD4 >500 after seroconversion, for those with SPVL ≥5.0 | 64-77 | Netherlands ATHENA cohort ^16^ |
| $f_{3,4}$ | Percentage with CD4 200-350 after seroconversion, for those with SPVL ≥5.0 | 2-7 | Netherlands ATHENA cohort ^16^ |
| $f_{4,4}$ | Percentage with CD4 <200 after seroconversion, for those with SPVL ≥5.0 | 0 (fixed) | Netherlands ATHENA cohort ^16^ |
| *Transmission probabilities* | |  |  |
| $d_{1}$ | Relative infectiousness of HIV-positive partner in acute stage of infection vs chronic & CD4>200 (off ART) | 4.47-18.81 | ^14^ |
| $d_{2}$ | Relative infectiousness of HIV-positive partner in late stage of infection – CD4<200 cells/µl vs chronic and CD4>200 (off ART) | 2-8 | ^24,25^ |
| $\beta$ | Average probability of acquiring HIV infection per sex act with an HIV-positive partner with chronic untreated infection | 0.0007-0.0285 | ^26,27^; assume 50% of sex acts are insertive |
| $h_{1}$ | Relative infectiousness of HIV-positive person with log_10_ SPVL <4.0 vs 4.0-4.5 | 0.337-0.68 | ^28^ Inverse of pooled increase in transmissibility per log10 decrease in viral load |
| $h_{2}$ | Relative infectiousness of HIV-positive person with log_10_ SPVL 4.0-4.5 vs 4.0-4.5 | 1 (fixed) |  |
| $h_{3}$ | Relative infectiousness of HIV-positive person with log_10_ SPVL 4.5-5.0 vs 4.0-4.5 | 1 (fixed) |  |
| $h_{4}$ | Relative infectiousness of HIV-positive person with log_10_ SPVL >5.0 vs 4.0-4.5 | 1.47-2.97 | ^28^ pooled increase in transmissibility per log10 increase in viral load |
| *Intervention behaviour* | |  |  |
| $p$ | Percentage of new entrants to MSM population who never routinely test for HIV | 5-13 | NHBS Baltimore MSM 2004-2011: % of those aged >24 years old who report never testing for HIV^1^ |
| $\tau_{0,0}$ | Percentage of undiagnosed black MSM aged 18-24 testing for HIV in the last year, 2004 onwards^a^ | 63.8-95.0 (reported testing rates)  25.5-47.5 (diagnosis fitting) | NHBS data 2004-2011, self-reported HIV negative men; converted into rate of testing at least once per year in the model^1^  Diagnosis fitting: 60% reduction |
| $\tau_{0,1}$ | Percentage of undiagnosed white MSM aged 18-24 testing for HIV in the last year, 2004 onwards^a^ | 32.1-82.3 (reported testing rates)  12.8-41.2 (diagnosis fitting) | NHBS data 2004-2008(highest and lowest from ranges), self-reported HIV negative men^1^  Diagnosis fitting: 60% reduction |
| $\tau_{1,0}$ | Percentage of undiagnosed black MSM aged >24 years old testing for HIV in the last year, 2004 onwards^a^ | 50.0-70.2 (reported testing rates)  20.0-35.1 (diagnosis fitting) | NHBS data 2004-2011 (highest and lowest from ranges), self-reported HIV negative men^1^  Diagnosis fitting: 60% reduction |
| $\tau_{1,1}$ | Percentage of undiagnosed white MSM aged >24 years old testing for HIV in the last year, 2004 onwards^a^ | 32.7-69.7 (reported testing rates)  13.1-34.9(diagnosis fitting) | NHBS data 2004-2011 (highest and lowest from ranges), self-reported HIV negative men^1^  Diagnosis fitting: 60% reduction |
| $\tau_{early}$ | Percentage of all MSM who tested for HIV in the last year, 1996 | 20-30 (reported testing rates)  8-15 (diagnosis fitting) | MSM in national NHSDA survey 1996 ^29^  Diagnosis fitting: 60% reduction |
| $\omega$ | Ratio of rate of dropout from care: rate of dropout from ART | 1-7 | Estimates from US studies - risk of dropout from care for those on vs off ART ^30-32^ |
| *q_1_* | Percentage of white MSM testing positive for HIV who link to care straight away | 72-86 | ^33-37^ |
| $\epsilon$ | Rate of linkage to care for those not linking immediately or dropped out, per year | 0-0.1 (fitting to care and viral suppression data) 0-0.2 0-0.5 (fitting to ART coverage data) | Estimate |
| linkage_inc | Annual absolute increase in percentage of white MSM who link to care straight away after testing positive for HIV | 3.5 (fixed) | From changes for MSM in national CDC data ^38,39^ |
| $\chi$ | Percentage of white MSM initiating ART who are adherent (achieve viral suppression) | 73-99 | ^37,40-42^ |
| $\xi_{x}$ | Rate of initiation onto ART from care, when meeting CD4 criteria^b^, per year^a^ | 0.5-2.1 (fitting to care and viral suppression data)  1.1-4 (fitting to ART coverage data) | Assuming CD4 testing every 3-6 months (national guidelines), acceptance 80-90% ^43^ |
| ${\psi_{0,z}, \psi}_{1,z}$ | Rate of starting HAART due to AIDS symptoms, CD4>500, per year (post-1996) | 0.002-0.01 | Incidence of AIDS-defining illness among ART naives, CASCADE collaboration ^44^; similar estimates from EURO-COORD data analysis ^45^ |
| $\psi_{2,z}$ | Rate of starting HAART due to AIDS symptoms, CD4 350-500, per year (post-1996) | 0.008-0.015 | Incidence of AIDS-defining illness among ART naives, CASCADE collaboration ^44^; similar estimates from EURO-COORD data analysis ^45^ |
| $\psi_{3,z}$ | Rate of starting HAART due to AIDS symptoms, CD4 200-350, per year (post-1996) | 0.018-0.032 | Incidence of AIDS-defining illness among ART naives, CASCADE collaboration ^44^; similar estimates from EURO-COORD data analysis ^45^ |
| $\psi_{4,z}$ | Rate of starting HAART due to AIDS symptoms, CD4<200, per year (post-1996) | 0.173-0.262 | Incidence of AIDS-defining illness among ART naives, CASCADE collaboration ^44^ |
| $\phi_{4}\phi_{5}\phi_{6}$ | Dropout from ART, not fully suppressed/1^st^ year on ART/2^nd^ year on ART, per year | 0.06-0.13 | Rate of dropout from ART, US ^46^ ^30^ ^31,32^,^47^ |
| $\phi_{z ratio}$ | Ratio of dropout from ART 3^rd^+ years: dropout 1^st^, 2^nd^ years ($\phi_{7}: \phi_{4}$) | 0.5-1.0 | Rate of dropout from US ART cohorts ^48^ |
| $\zeta$ | Rate of re-enrolment into pre-ART HIV care for those dropping out of ART, per year | 0.05-1 | From rate of dropout and re-joining US ART cohorts ^48^ |
| $s_{n}$ | Percentage of MSM circumcised | 77-89 | NHBS 2008 & 2011 ^1^ |
| $\epsilon_{ratio}$ | Ratio of rates of linkage to care for black:white MSM (ratio also applied to percentage linking immediately after diagnosis) | 1-2 (fitting to care and viral suppression data)  0.84-1.5 (fitting to ART coverage data) | ^33,37^ |
| $\omega_{ratio}$ | Ratio of dropout from care for white:black MSM | 1-3 (fitting to care and viral suppression data)  0.46-1.54 (fitting to ART coverage data) | ^30,46,47^ ^31,32,49^ |
| $\xi_{ratio}$ | Ratio of ART initiation rate for black:white MSM | 0.4-1.0 | ^32^ |
| $\phi_{w ratio}$ | Ratio of ART dropout for black:white MSM | 0.7-1.6 | ^46,48^ |
| $\chi_{ratio}$ | Ratio of percentage adherent to ART black:white MSM | 0.82-1 | ^30,37,40-42,50^ |
| $\delta$ | Proportion of negative HIV tests after which PrEP is offered in 2020 | 0.08-0.354 | Range explored; rate increases linearly from 0 in 2012 |
| $s_{p00}$ | Adherence to PrEP (% taking ≥4 doses/week), 18-24 year old black MSM | 0.63 (fixed) | Stratified analysis of US PrEP Demo project data among those clinic-referred |
| $s_{p01}$ | Adherence to PrEP (% taking ≥4 doses/week), 18-24 year old white MSM | 0.9 (fixed) | Stratified analysis of US PrEP Demo project data among those clinic-referred |
| $s_{p10}$ | Adherence to PrEP (% taking ≥4 doses/week), >24 year-old black MSM | 0.42 (fixed) | Stratified analysis of US PrEP Demo project data among those clinic-referred |
| $s_{p11}$ | Adherence to PrEP (% taking ≥4 doses/week), >24 year-old white MSM | 0.87 (fixed) | Stratified analysis of US PrEP Demo project data among those clinic-referred |
| $\iota_{00}$ | PrEP dropout, 18-24 year old black MSM | 0.67 (fixed) | Proportion not retained at the end of the study, stratified analysis of US PrEP Demo project data among those clinic-referred |
| $\iota_{01}$ | PrEP dropout, 18-24 year old white MSM | 0.25 (fixed) | Proportion not retained at the end of the study, stratified analysis of US PrEP Demo project data among those clinic-referred |
| $\iota_{10}$ | PrEP dropout >24 year-old black MSM | 0.19 (fixed) | Proportion not retained at the end of the study, stratified analysis of US PrEP Demo project data among those clinic-referred |
| $\iota_{11}$ | PrEP dropout >24 year-old white MSM | 0.21 (fixed) | Proportion not retained at the end of the study, stratified analysis of US PrEP Demo project data among those clinic-referred |
| *Intervention efficacy* | |  |  |
| $e_{c}$ | Per-sex-act reduction in HIV acquisition risk due to correct condom use (%) | 58-79 | Estimate for US MSM ^51^ |
| $e_{n}$ | Per-sex-act reduction in HIV acquisition risk due to male circumcision (%) | 12-23 | Assuming same efficacy as for heterosexual men from RCTs ^52^, only protective in insertive acts, half of all sex acts are insertive, receptive sex acts carry a 2.3x higher risk of transmission than insertive^27^. |
| $d_{r}$ | Relative level of infectiousness of those on ART and partially suppressed, scaled between the level for those fully suppressed ${(d}_{r}=0)$.and those unsuppressed ($d_{r}=1)$ | 0.5(fixed) | assumption |
| $d_{6}$ | Per-sex-act reduction in HIV transmission risk when on ART and fully suppressed vs chronic infection untreated (CD4>200) (%) | 99-100 | Estimates from discordant MSM partnerships where HIV-positive partner on ART and virally suppressed ^53^ |
| $e_{p}$ | Per-sex-act reduction in HIV acquisition risk when adherent to PrEP (taking ≥4 tablets/week) | 90 (fixed) | Minimum efficacy estimated from iPrEx and STRAND trial data analysis for 4 doses/week^54^ |

^a^Final values for time-varying parameters. Earlier values or earlier gradient of parameter function given elsewhere in table S1.

^b^Guideline changes coded in: pre-1996, no initiation of ART ^55^ From 1996-1998 ART initiation at any CD4 count; from 1998-Feb 2001, initiation from care with CD4<500 (1998 guidelines); from Feb 2001-Dec 2009 initiation with CD4 <350 (2001 guidelines); from Dec 2009-March 2012 initiation from care with CD4<500 (2009 guidelines); from March 2012 onwards initiation from care with any CD4 count (2012 guidelines). These apply to all age and race groups.

**Table S2: Data fitted to, with fitting bounds, source and justification**

| **Output** | **Year** | **Estimate** | **Min** | **Max** | **Source & justification** | **Fitting assumption used for** | | **Used for validation** |
| --- | --- | --- | --- | --- | --- | --- | --- | --- |
| **Demography** |  |  |  |  |  | NHBS age/race distribution | Census age/race distribution |  |
| Total MSM population size | 2010 | 6518 | 4270 | 8765 | Range 1.9-3.9%^7^ of male population aged 18+ in Baltimore 2010 census (224,742) | ✓ | ✓ |  |
| Percentage of population aged 18-24 | 1990 | 15 | 10 | 20 | Census estimate ± 5pp |  | ✓ |  |
|  | 2000 | 15 | 10 | 20 | Census estimate ± 5pp |  | ✓ |  |
|  | 2010 | 16 | 11 | 21 | Census estimate ± 5pp |  | ✓ |  |
|  | 2004 | 24.8 | 20.3 | 30.0 | NHBS data 95% CI^1^ | ✓ |  |  |
|  | 2008 | 30.6 | 22.9 | 40.0 | NHBS data 95% CI^1^ | ✓ |  |  |
|  | 2011 | 30.9 | 21.5 | 42.2 | NHBS data 95% CI^1^ | ✓ |  |  |
|  | 2014 | 23.9 | 18.0 | 31.0 | NHBS data 95% CI^1^ |  |  | ✓ |
| Percentage of white MSM aged 18-24 | 2010 | 14 | 9 | 19 | Census estimate ± 5pp |  | ✓ |  |
|  | 2004 | 21.0 | 15.6 | 27.6 | NHBS data 95% CI^1^ | ✓ |  |  |
|  | 2008 | 17.1 | 9.4 | 29.3 | NHBS data 95% CI^1^ | ✓ |  |  |
|  | 2011 | 20.7 | 11.8 | 33.7 | NHBS data 95% CI^1^ | ✓ |  |  |
|  | 2014 | 14.6 | 8.6 | 23.6 | NHBS data 95% CI^1^ |  |  | ✓ |
| Percentage of black MSM aged 18-24 | 2010 | 16 | 11 | 21 | Census estimate ± 5pp |  | ✓ |  |
|  | 2004 | 24.0 | 18.0 | 31.2 | NHBS data 95% CI^1^ | ✓ |  |  |
|  | 2008 | 32.5 | 23.8 | 42.6 | NHBS data 95% CI^1^ | ✓ |  |  |
|  | 2011 | 34.3 | 22.3 | 48.7 | NHBS data 95% CI^1^ | ✓ |  |  |
|  | 2014 | 27.2 | 19.4 | 36.7 | NHBS data 95% CI^1^ |  |  | ✓ |
| Percentage of MSM who are black | 1990 | 61 | 56 | 66 | Census estimate ± 5pp |  | ✓ |  |
|  | 2000 | 68 | 63 | 73 | Census estimate ± 5pp |  | ✓ |  |
|  | 2010 | 69 | 64 | 74 | Census estimate ± 5pp |  | ✓ |  |
|  | 2004 | 64.1 | 49.8 | 76.3 | NHBS data 95% CI^1^ | ✓ |  |  |
|  | 2008 | 73.1 | 59.3 | 83.6 | NHBS data 95% CI^1^ | ✓ |  |  |
|  | 2011 | 84.2 | 71.6 | 91.8 | NHBS data 95% CI^1^ | ✓ |  |  |
|  | 2014 | 73.8 | 64.6 | 81.3 | NHBS data 95% CI^1^ |  |  | ✓ |
| **HIV prevalence** |  |  |  |  |  | All fitting assumptions |  |  |
| HIV prevalence black MSM aged 18-24 years old | 2004 | 33.0 | 23.6 | 43.8 | NHBS data 95% CI^1^ | ✓ |  |  |
|  | 2008 | 31.2 | 20.6 | 44.2 | NHBS data 95% CI^1^ | ✓ |  |  |
|  | 2011 | 39.6 | 32.0 | 47.8 | NHBS data 95% CI^1^ | ✓ |  |  |
|  | 2014 | 24.1 | 14.5 | 37.1 | NHBS data 95% CI^1^ |  |  | ✓ |
| HIV prevalence black MSM aged >24 years old | 2004 | 58.4 | 46.7 | 69.3 | NHBS data 95% CI^1^ | ✓ |  |  |
|  | 2008 | 51.8 | 42.9 | 60.7 | NHBS data 95% CI^1^ | ✓ |  |  |
|  | 2011 | 52.2 | 44.1 | 60.3 | NHBS data 95% CI^1^ | ✓ |  |  |
|  | 2014 | 47.4 | 39.4 | 55.5 | NHBS data 95% CI^1^ |  |  | ✓ |
| HIV prevalence white MSM aged 18-24 years old | 2004 |  | 0 | 100 | Numbers too small^1^ |  |  |  |
|  | 2008 | 22.2 | 10.1 | 42.0 | NHBS data 95% CI^1^ | ✓ |  |  |
|  | 2011 |  | 0 | 100 | Numbers too small^1^ |  |  |  |
|  | 2014 |  | 0 | 100 | Numbers too small^1^ |  |  |  |
| HIV prevalence white MSM aged >24 years old | 2004 | 16.7 | 10.7 | 25.0 | NHBS data 95% CI^1^ | ✓ |  |  |
|  | 2008 | 18.4 | 9.4 | 32.9 | NHBS data 95% CI^1^ | ✓ |  |  |
|  | 2011 | 19.6 | 12.2 | 29.8 | NHBS data 95% CI^1^ | ✓ |  |  |
|  | 2014 | 9.1 | 5.0 | 15.9 | NHBS data 95% CI^1^ |  |  | ✓ |
| **Care continuum indicators** |  |  |  |  |  | NHBS HIV testing rate | CDC estimates for Maryland |  |
| Percentage of HIV-positive MSM diagnosed | 2012 | 75.9 | 71.7 | 80.5 | CDC data for Maryland state^56^95% CI |  | ✓ |  |
|  |  |  |  |  |  | NHBS ART coverage data | Maryland DH continuum data |  |
| Percentage of all HIV-positive MSM on ART | 2008 | 39.5 | 31.9 | 47.5 | NHBS ARV detection analysis ^57^95% CI^1^ | ✓ |  |  |
|  | 2011 | 55.4 | 48.0 | 62.6 | NHBS ARV detection analysis 95% CI^1^ | ✓ |  |  |
|  | 2014 | 70.3 | 61.6 | 77.7 | NHBS ARV detection analysis 95% CI^1^ |  |  | ✓ |
| Percentage of black HIV-positive MSM on ART | 2008 | 36.9 | 28.5 | 46.2 | NHBS ARV detection analysis ^57^95% CI^1^ | ✓ |  |  |
|  | 2011 | 51.6 | 43.8 | 59.4 | NHBS ARV detection analysis 95% CI^1^ | ✓ |  |  |
|  | 2014 | 70.2 | 60.8 | 78.1 | NHBS ARV detection analysis 95% CI^1^ |  |  | ✓ |
| Percentage of white HIV-positive MSM on ART | 2008 | 61.1 | 38.6 | 79.7 | NHBS ARV detection analysis ^57^95% CI^1^ | ✓ |  |  |
|  | 2011 |  | 0 | 100 | Numbers too small^1^ |  |  |  |
|  | 2014 |  | 0 | 100 | Numbers too small^1^ |  |  |  |
| Percentage of diagnosed black MSM in care | 2012-2013 | 63.5 | 56.7 | 70.3 | Maryland DH^a^ ± 5pp min-max for 2012-2013^1^ |  | ✓ |  |
|  | 2014 | 63.1 | 58.1 | 68.1 | Maryland DH^a^ ± 5pp^1^ |  |  | ✓ |
|  | 2015 | 57.3 | 52.3 | 62.3 | Maryland DH^a^ ± 5pp^1^ |  |  | ✓ |
| Percentage of diagnosed white MSM in care | 2012-2013 | 51.6 | 44.6 | 58.6 | Maryland DH^a^ ± 5pp min-max for 2012-2013^1^ |  | ✓ |  |
|  | 2014 | 54.1 | 49.1 | 59.1 | Maryland DH^a^ ± 5pp^1^ |  |  | ✓ |
|  | 2015 | 44.8 | 39.8 | 49.8 | Maryland DH^a^ ± 5pp^1^ |  |  | ✓ |
| Percentage of diagnosed black MSM virally suppressed | 2012 | 31.6 | 26.6 | 36.6 | Maryland DH^b^ ± 5pp^1^ |  | ✓ |  |
|  | 2013 | 37.0 | 32.0 | 42.0 | Maryland DH^b^ ± 5pp^1^ |  | ✓ |  |
|  | 2014 | 41.1 | 36.1 | 46.1 | Maryland DH^b^ ± 5pp^1^ |  |  | ✓ |
|  | 2015 | 39.9 | 34.9 | 44.9 | Maryland DH^b^ ± 5pp^1^ |  |  | ✓ |
| Percentage of diagnosed white MSM virally suppressed | 2012 | 35.1 | 30.1 | 40.1 | Maryland DH^b^ ± 5pp^1^ |  | ✓ |  |
|  | 2013 | 38.5 | 33.5 | 43.5 | Maryland DH^b^ ± 5pp^1^ |  | ✓ |  |
|  | 2014 | 38.6 | 33.6 | 43.6 | Maryland DH^b^ ± 5pp^1^ |  |  | ✓ |
|  | 2015 | 37.6 | 32.6 | 42.6 | Maryland DH^b^ ± 5pp^1^ |  |  | ✓ |
| Percentage of MSM on ART virally suppressed | 2010 | 85 | 75 | 90 | National estimates for MSM^37,58^ range | ✓ | ✓ |  |
| **PrEP coverage** |  |  |  |  |  | NHBS data |  |  |
| Percentage of all MSM taking PrEP | 2014 |  | 1.1 | 2.8 | Upper bound: NHBS data 2014, use in last 12 months^5^; lower bound 40% of this | ✓ |  |  |
|  | 2017 |  | 4.9 | 12.3 | Upper bound: NHBS data 2017, use in last 12 months^6^; lower bound 40% of this | ✓ |  |  |

^a^definition of in care: percentage of those diagnosed with at least one CD4 test past 12 months

^b^definition of virally suppressed: percentage of those diagnosed with at least one viral load test last 12 months and most recent viral load <200 copies/ml

**MODEL FITS TO DATA**

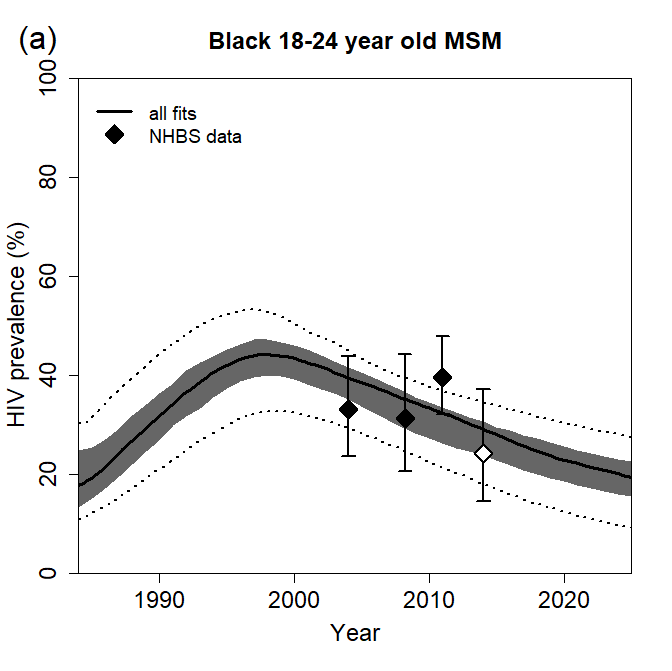

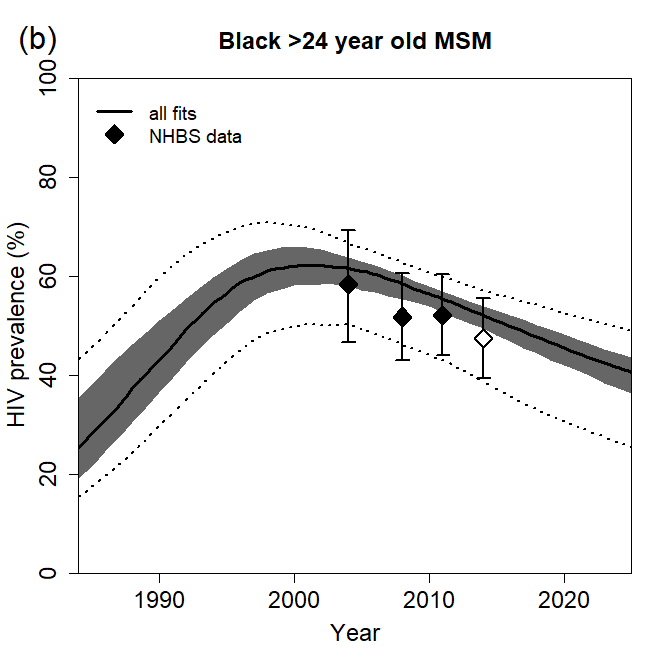

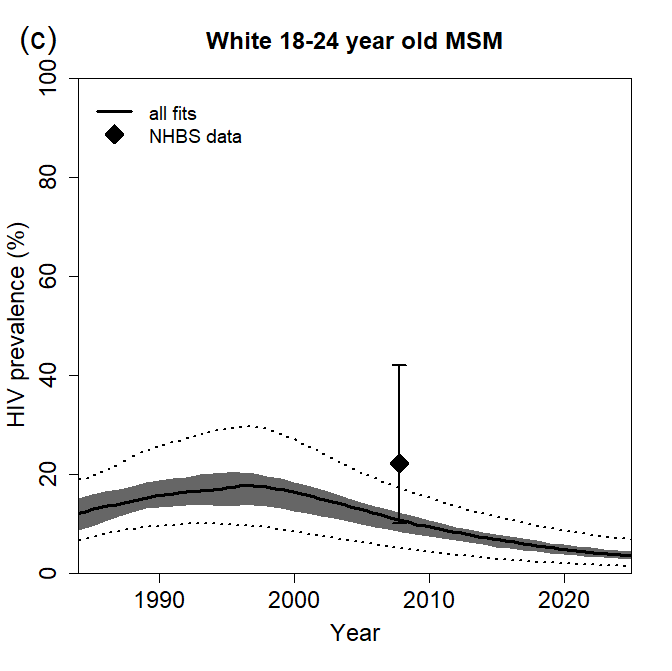

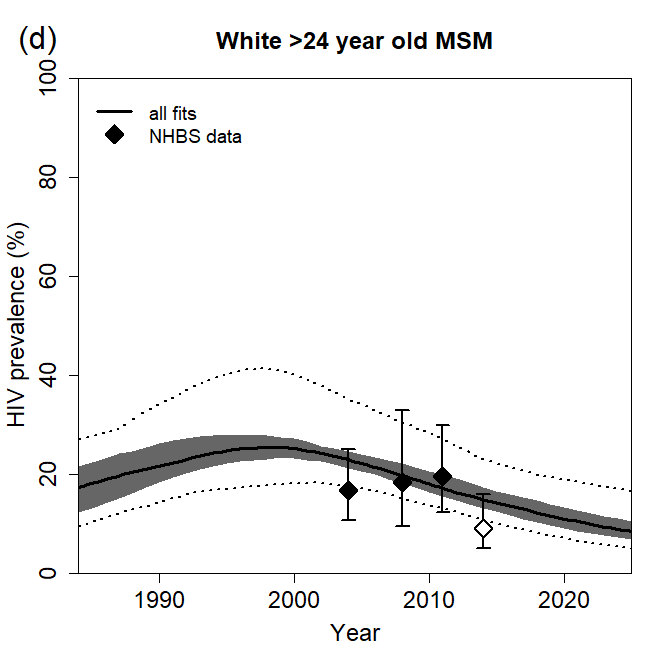

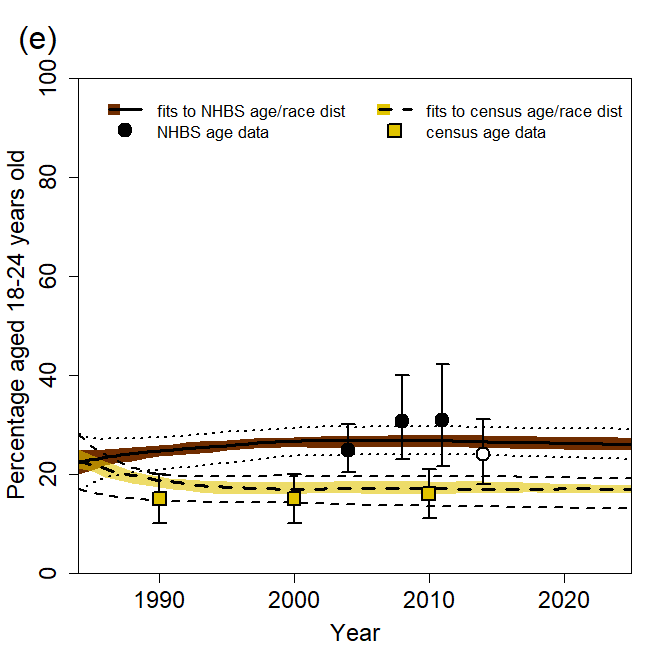

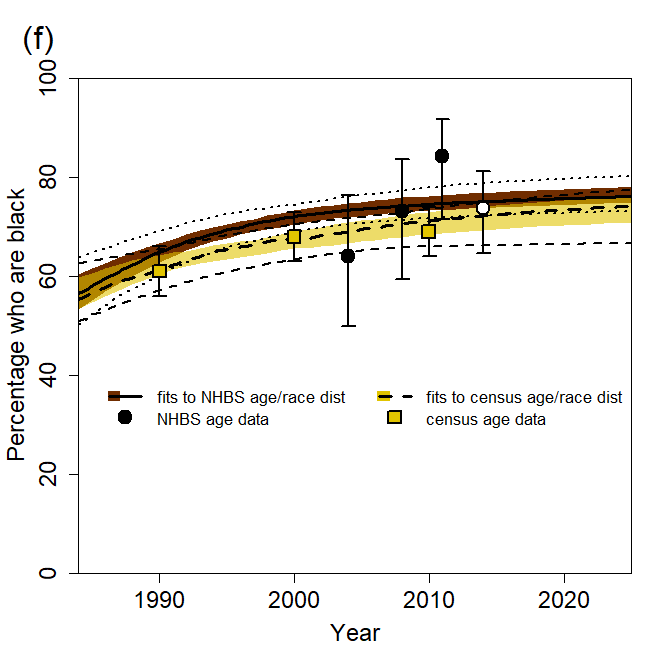

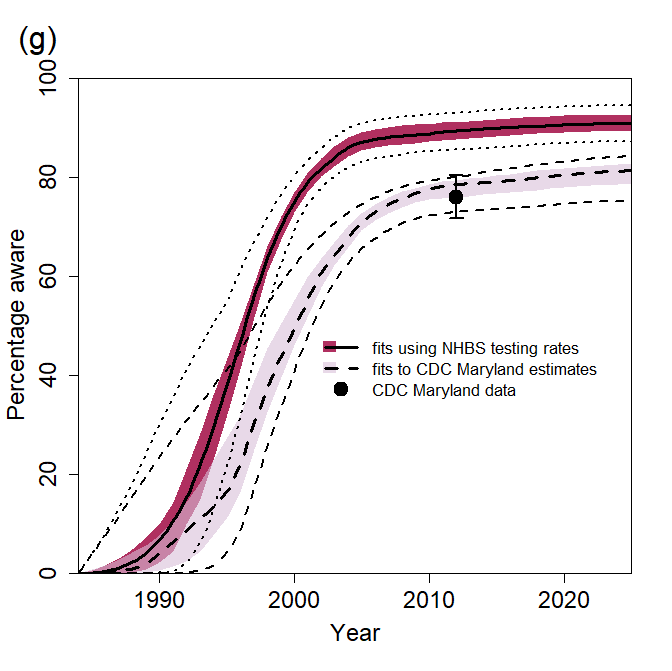

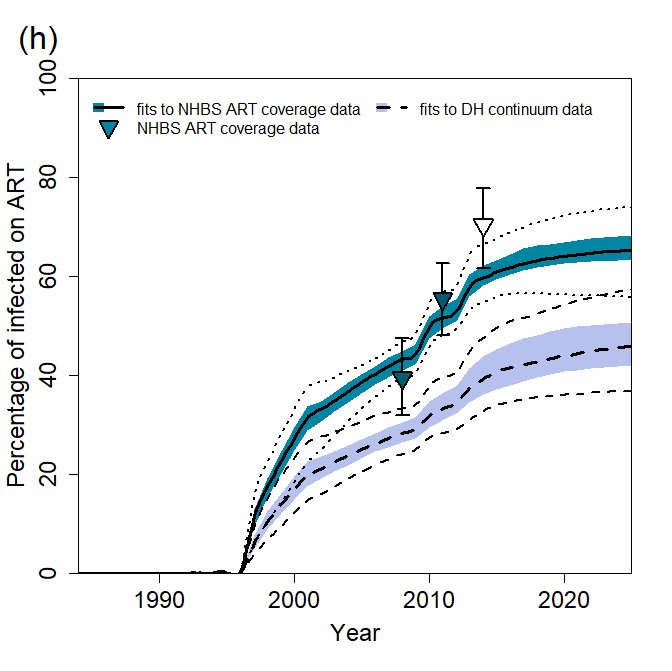

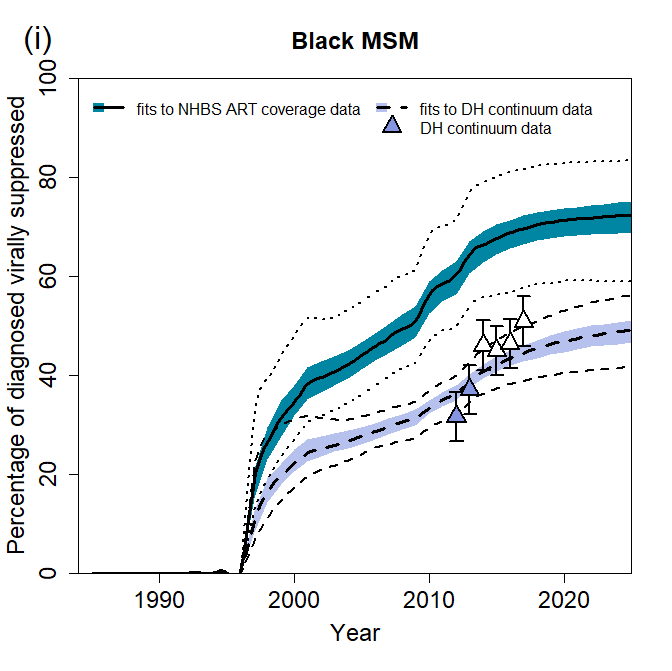

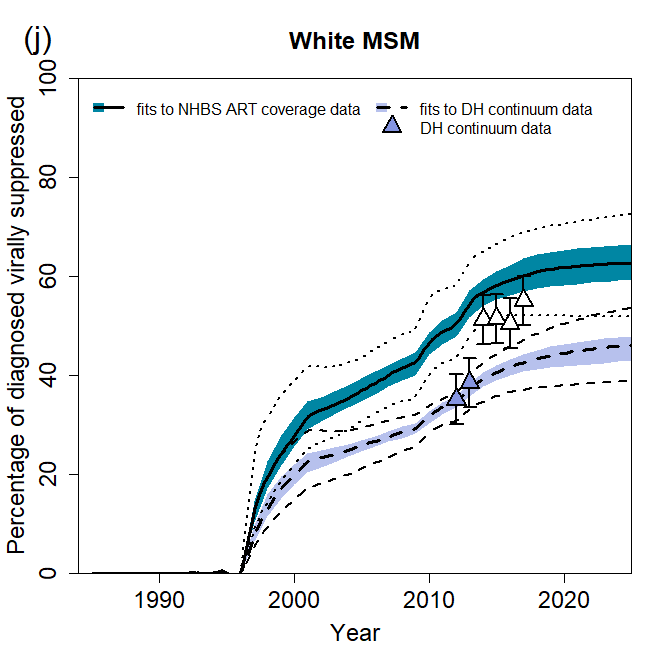

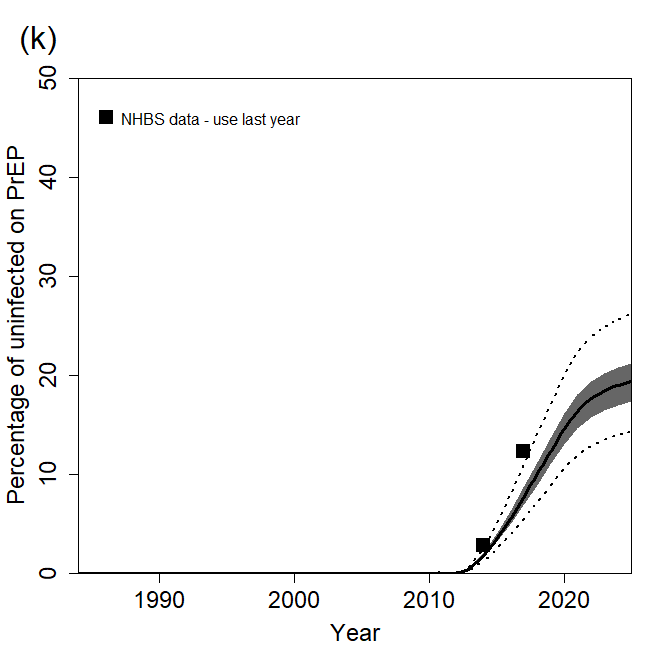

**Figure S4. Model fits to available data for MSM in Baltimore.** (a-d) HIV prevalence among young (18-24 year old) black/older (>24 year old) black/young white/older white MSM, (e) percentage of all MSM aged 18-24 years old, (f) percentage of MSM who are black, (g) percentage of HIV-positive MSM who are aware of their HIV-positive status, (h) percentage of all HIV-positive MSM who are on ART, (i) percentage of black diagnosed HIV-positive MSM who are virally suppressed, (j) percentage of white diagnosed HIV-positive MSM who are virally suppressed, (k) percentage of uninfected MSM who are taking PrEP. Results are for all 169 fitting parameter combinations. Results show median (thick lines), 25^th^-75^th^ percentile (dark shaded area), and 2.5^th^ and 97.5^th^ percentiles (dotted lines) across model fits. Points and error bars show the mean and 95% CI for National HIV Behavioural Surveillance (NHBS) data (a-f,h), mean and ±5 percentage points for census data (e,f) and DH continuum data (i,j), or mean for NHBS data (k). Data prior to 2014 (filled points) were used for model fitting. Data from 2014 (unfilled points) were used to validate model predictions. Number of fits under each assumption: demography fitting assumptions, NHBS age/race distribution (N=146), census age/race distribution (N=23); diagnosis fitting assumptions, NHBS HIV testing rate parameter (N=118), CDC estimates for Maryland (N=51); continuum fitting assumptions, NHBS ART coverage data (N= 101), DH continuum data (N= 68).

**Table S3: Magnitude of disruptions modelled, with source and justification**

| **Disruption to:** | **Overall data-driven reduction used in main scenario** | **Age- and/or race-specific reductions used in main scenario** | **Overall values explored in sensitivity analysis (age-/race-specific estimates adjusted proportionately)** | **Source/justification** |
| --- | --- | --- | --- | --- |
| HIV testing | 20% | 18-24-year-olds: 25%  ≥25-year-olds: 19% | 50%, 75% , 100% | Main estimate from Sanchez et al survey of US MSM.^59^ 18.8% report decreased access to HIV testing; 18.1% (52/286) of those who tried to get an HIV test had trouble getting one. Consistent with data from Stephenson et al survey of US MSM^60^: 32.2% reported that COVID-19 prevented them from getting a test for HIV. Age-specific reductions based on ratios from Sanchez et al^59^ |
| ART initiations | 50% | ·· | 50%, 75%, 100% | Assumption (no data found) |
| Viral suppression | 10% | White: 9%  Black: 15% | 10%, 25%, 50% | Main estimate from Sanchez et al survey of US MSM.^59^ Of those living with HIV, 24% report having fewer viral load or other lab tests, 6% report reduced access to ART medications, 9.5% (10/105) of those who’ve tried to get an ART prescription report trouble getting one, 5% report that they are taking their medications daily less often.  Race-specific reduction based upon data from Santos et al global survey of MSM,^61^ using ratio for those identifying as racial minority vs. not racial minority in % who either cannot refill/access ART or can refill/access ART with complications. ^61^ |
| PrEP initiations | 72% | ·· | 50%, 75%, 100% | Krakower et al study at a Boston PrEP clinic.^62^ 72% reduction in PrEP initiations in April 2020 vs January 2020. |
| PrEP use | 9% | Black 18-24-yr-olds: 13%  White 18-24-yr-olds: 11%  Black ≥25-yr-olds: 9%  White ≥25-yr-olds: 8% | 10%, 25%, 50% | Krakower et al study at a Boston PrEP clinic^62^: 9.2% excess PrEP lapses in April 2020 vs January 2020; in January, 4.4% (140/3197) lapsed, in April 13.6% (407/2984) lapsed.  Sanchez et al survey of US MSM ^59^: 12.9% (18/158) of those who tried to get a PrEP prescription had trouble getting one, and 8% (12/150) of those who tried to get PrEP medication had trouble getting it.  Stephenson et al survey of US MSM^60^: 8.9% report that COVID-19 has prevented access to a PrEP prescription.  Age and race differences estimated from ratios of % lapsed by age and race during lockdown at a Boston PrEP clinic^62^: 18.0% of under-26 years-olds and 12.6% of 27+ year-olds had a PrEP lapse in April 2020; 14.2% of Black patients and 12.4% of White patients had a PrEP lapse in April 2020. |
| HIV testing on PrEP | 85% | ·· | 50%, 75%, 100% | Krakower et al study at a Boston PrEP clinic.^62^ 85% reduction in number of HIV tests among those on PrEP in April 2020 vs January 2020. |
| Condom use | 5% | ·· | 10%, 25%, 50% | Sanchez et al survey of US MSM.^59^ 5.8% reported less condom use, 0.4% reported more condom use. N.B. Small differences in condom use by age reported by Sanchez et al^59^ not explored as the model does not allow for condom use differing by age. |
| Partner numbers | 25%, 50% | 18-24-year olds: 22%, 44%  ≥25-year-olds: 26%, 52% | 10%, 25%, 50% | Sanchez et al survey of US MSM^59^ 51.3% reported having fewer sex partners, 0.8% reported having more. McKay et al survey of US MSM (9) overall 39.5% reported having fewer sex partners, 1.2% reported having more.  Age-specific reductions based on ratios from Sanchez et al.^59^ |

**Supplementary Results**

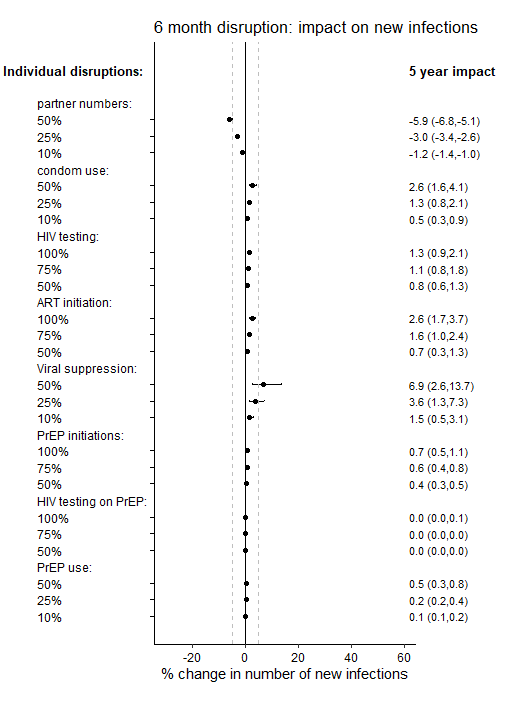

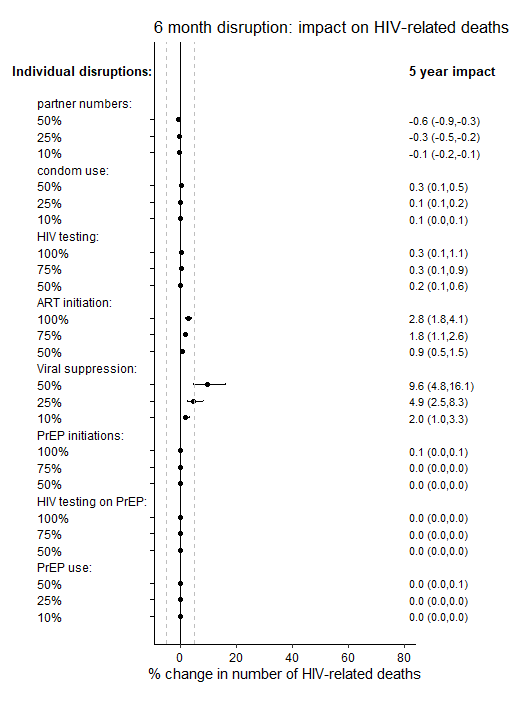

**(a)**

**(b)**

**Figure S5: Sensitivity analysis on disruption magnitude - 5-year impact.** Impact of individual disruptions due to COVID-19, exploring different values for the size of these disruptions, indicated on plot. Impact on (a) cumulative new HIV infections and (b) cumulative HIV-related deaths, over 5 years. Points are median and error bars are 95% credible intervals across all model fits. Disruptions are assumed to last for 6 months. Dashed vertical lines are at -5% and 5% .

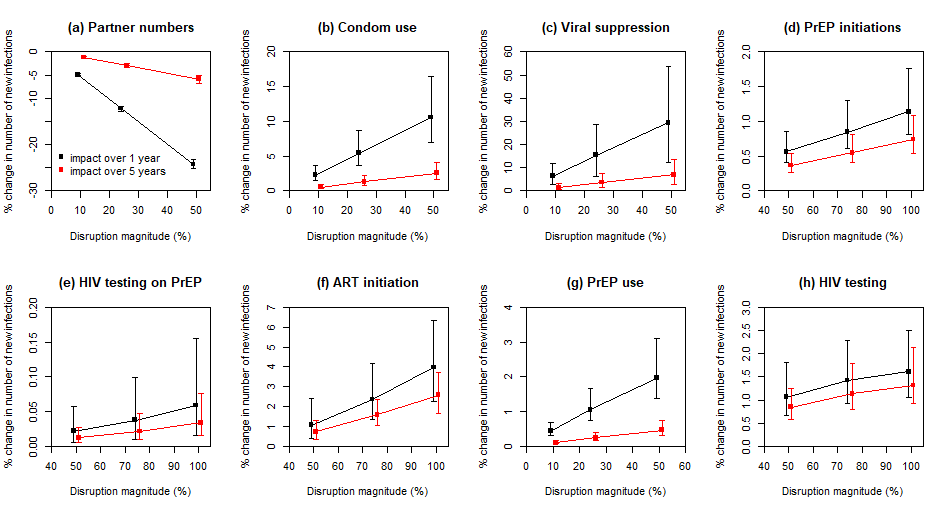

**Figure S6: Sensitivity analysis on disruption magnitude: impact on new HIV infections.** Impact of individual disruptions due to COVID-19, exploring different values for the size of these disruptions, indicated on plot. Impact on cumulative new HIV infections over 1 year (black points) and 5 years (red points). Points are median and error bars are 95% credible intervals across all model fits. Disruptions are assumed to last for 6 months.

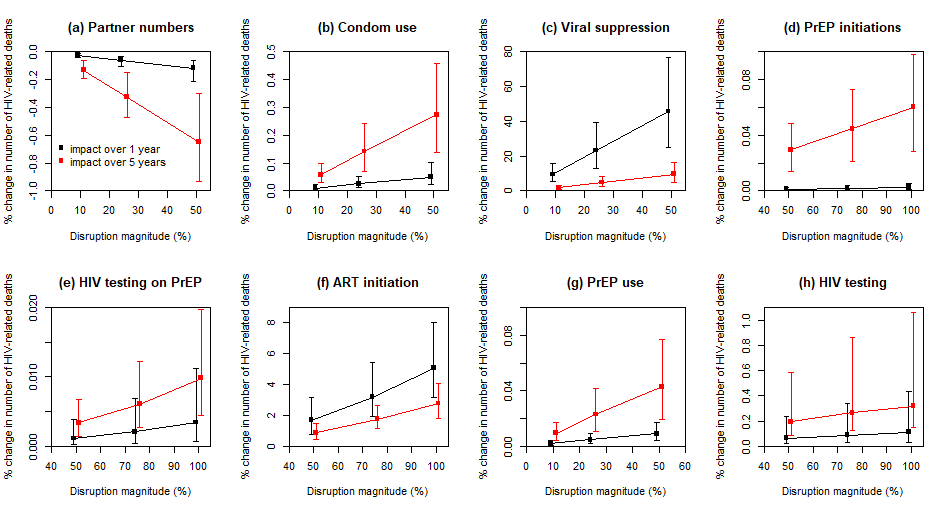

**Figure S7: Sensitivity analysis on disruption magnitude: impact on HIV-related deaths.** Impact of individual disruptions due to COVID-19, exploring different values for the size of these disruptions, indicated on plot. Impact on HIV-related deaths over 1 year (black points) and 5 years (red points). Points are median and error bars are 95% credible intervals across all model fits. Disruptions are assumed to last for 6 months.

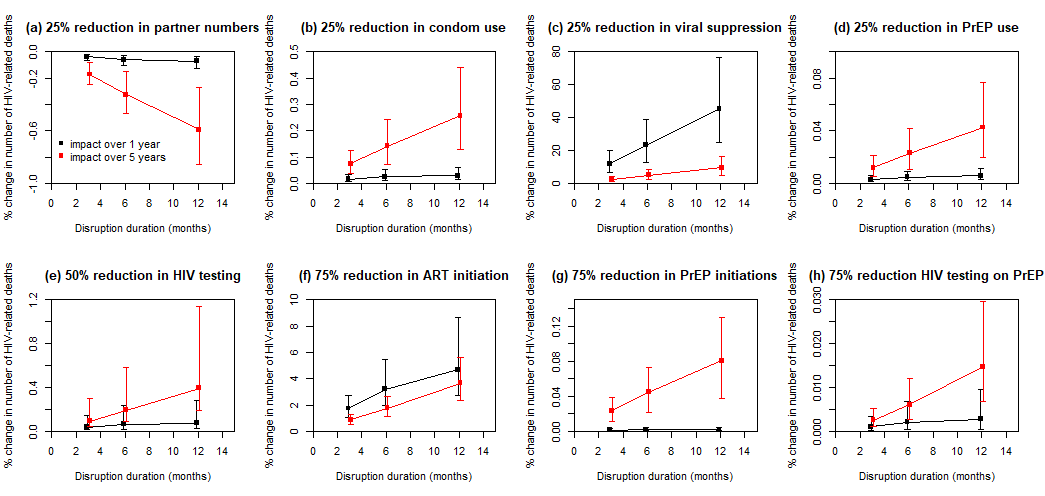

**Figure S8. Impact of disruptions lasting 3, 6 or 12 months on HIV-related deaths.** Impact on cumulative HIV-related deaths over 1 year (black points) and 5 years (red points), for disruptions indicated. Points are median and error bars are 95% credible intervals across all model fits. Note very different y-axis scales used to clearly show linearity of trends.

**Disruption to partner numbers only**

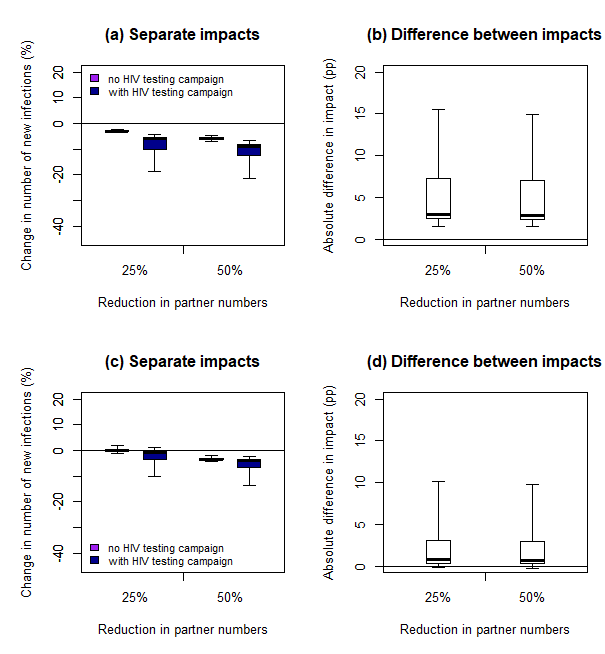

**Disruption to HIV testing, ART, PrEP, condom use and partner numbers**

**Figure S9.** **Impact of disruptions with and without additional HIV testing campaign, measured over 5 years.** Impact on cumulative new HIV infections over five years of six-month disruptions to (a,b) partner numbers only or (c,d) HIV testing, ART initiation, viral suppression, PrEP initiation and continuation and condom use as well as partner numbers, with (dark blue bars) or without (purple bars) an additional HIV testing campaign reaching 90% of MSM for HIV testing during the six-month disruption. For the full disruption (c,d), 20% reduction in HIV tests, 5% reduction in condom use , 72% reduction in PrEP initiations, 40% reduction in PrEP refills, 85% reduction in HIV testing on PrEP, 50% reduction in new ART initiations and 10% reduction in viral suppression (see table 1 for age- and race-specific disruptions). Thick lines are median, boxes are interquartile range, and whiskers full range across all model fits.

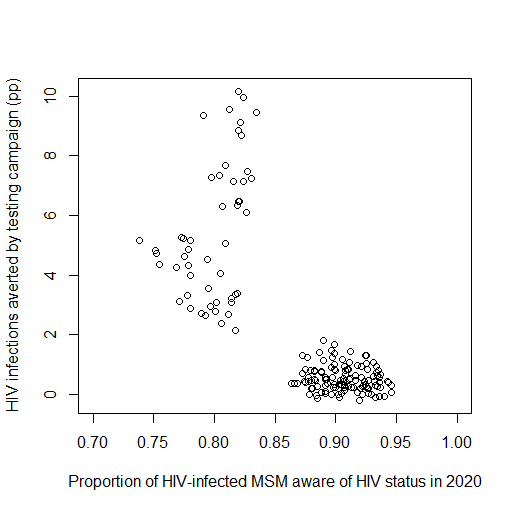

**Figure S10. Absolute impact of the HIV testing campaign on new HIV infections plotted against levels of awareness of HIV-positive status in 2020.** Data from Figure 4d (with 25% reduction in partnerships) are replotted against awareness levels. The absolute impact is the absolute difference between the impact of a scenario without an HIV testing campaign during the period of disruption and the impact of a scenario with the testing campaign, expressed as percentage points (c.f. Figure 4b,d). Disruptions are assumed to last for 6 months and consist of: a 20% reduction in HIV tests, 5% reduction in condom use , 72% reduction in PrEP initiations, 40% reduction in PrEP refills, 85% reduction in HIV testing on PrEP, 50% reduction in new ART initiations, 10% reduction in viral suppression and 25% reduction in partner numbers(see table 1 for age- and race-specific disruptions). The HIV testing campaign reaches 90% of MSM for HIV testing during the six-month disruption. Impact is measured over 1 year. Points are from individual model fits.

34.	Center for HIV Surveillance Epidemiology and Evaluation Maryland Department of Health and Mental Hygiene. Baltimore City annual HIV epidemiological profile 2013. Baltimore, MD. , 2015.

35.	Center for HIV Surveillance Epidemiology and Evaluation Maryland Department of Health and Mental Hygiene. 2012 Baltimore City annual HIV epidemiological profile. Baltimore, MD. , 2015.

36.	Center for HIV Surveillance Epidemiology and Evaluation Maryland Department of Health and Mental Hygiene. Baltimore City annual HIV epidemiological profile 2015. 2016.

62.	Krakower D, Solleveld P, Levine K, Mayer K. Impact of COVID-19 on HIV preexposure prophylaxis care at a Boston community health center [abstract number OACLB0104]. 23rd International AIDS conference. virtual; 2020.
